# The tryptophan catabolite or kynurenine pathway in a major depressive episode with melancholia, psychotic features and suicidal behaviors; a systematic review and meta-analysis

**DOI:** 10.1101/2022.08.06.22278492

**Authors:** Abbas F. Almulla, Yanin Thipakorn, Asara Vasupanrajit, Chavit Tunvirachaisakul, Gregory Oxenkrug, Hussein K. Al-Hakeim, Michael Maes

## Abstract

**Background:** Major depressive disorder (MDD) and bipolar disorder (BD) with melancholia and psychotic features and suicidal behaviors are accompanied by activated immune-inflammatory and oxidative pathways which may stimulate indoleamine 2,3- dioxygenase (IDO), the first and rate-limiting enzyme of the tryptophan catabolite (TRYCAT) pathway resulting in increased tryptophan degradation and elevated tryptophan catabolites (TRYCTAs).

**Objective:** The purpose of the current study is to systematically review and meta-analyze levels of TRP, its competing amino-acids (CAAs) and TRYCATs in patients with severe affective disorders.

**Methods:** PubMed, Google Scholar and SciFinder were searched in the present study and we recruited 35 studies to examine 4,647 participants including 2,332 unipolar (MDD) and bipolar (BD) depressed patients and 2,315 healthy controls.

**Results:** Severe patients showed significant lower (p<0.0001) TRP (standardized mean difference, SMD=-0.517, 95% confidence interval, CI: -0.735; -0.299) and TRP/CAA (SMD= -0.617, CI: -0.957; -0.277) levels with moderate effect sizes, while no significant difference in CAAs were found. Kynurenine (KYN) levels were unaltered in severe MDD/BD phenotypes, while the KYN/TRP ratio showed a significant increase only in patients with psychotic features (SMD= 0.224, CI: 0.012; 0.436). Quinolinic acid (QA) was significantly increased (SMD= 0.358, CI: 0.015; 0.701) and kynurenic acid (KA) significantly decreased (SMD= -0.260, CI: -0.487; -0.034) in severe MDD/BD.

**Conclusion:** Patients with affective disorders with melancholic and psychotic features and suicidal behaviors show normal IDO enzyme activity but a lowered availability of plasma/serum TRP to the brain, which is probably due to other processes such as low albumin levels.

## Introduction

Major depression disorder (MDD) and bipolar disorder (BD) may involve severe phenotypes including melancholia and psychotic depression (American Psychiatric Association 2000). Delusions and hallucinations are the main characteristics of the psychotic features in MDD, while melancholic MDD is characterized by severe depressed mood, anhedonia, hypoesthesia, lack of reactivity, early morning awakening, diurnal variation, and anorexia resulting in weight loss, and psychomotor retardation. These subtypes of MDD are strongly associated with suicidal behaviors in MDD patients (Sowa-Kućma, Styczeń et al. 2018). Suicide is one of the major causes of death worldwide, one of each 100 deaths is due to suicide, and globally 800,000 individuals die per year (World Health 2019, World Health 2021). The presence of psychiatric illness, particularly MDD and BD, is the major leading cause of suicide, particularly MDD (around 90% of the victims) (Henriksson, Aro et al. 1993, Lesage, Boyer et al. 1994).

Extensive evidence is now available indicating the involvement of activated immune-inflammatory and oxidative and nitrosative stress pathways (IO&NS) in the pathophysiology of mental disorders including MDD, BD and schizophrenia (Maes, Galecki et al. 2011, Martin-Subero, Anderson et al. 2016, Maes, Sirivichayakul et al. 2020, Solmi, Suresh Sharma et al. 2021, Vasupanrajit, Jirakran et al. 2022). Moreover, MDD and BD are accompanied by activation of the immune-inflammatory response system (IRS) reflected by alterations of acute phase proteins (APPs) (e.g. C-reactive protein and albumin) and activation of cell-mediated immunity as shown by increased interleukin (IL)- 6, tumor necrosis factor (TNF)-α, IL-1β, IL-2, interferon (IFN)-γ, soluble IL-2 receptor (sIL-2R), sCD8, high levels of activated T cells such as CD25+ and HLA-DR+) (Vasupanrajit, Jirakran et al. 2022). IRS is usually counterbalanced by the compensatory immune-regulatory system (CIRS), which increases T-regulatory cytokines such as IL-10 and transforming growth factor (TGF)-β (Maes, Galecki et al. 2011, Maes and Carvalho 2018, Roomruangwong, Noto et al. 2020, Maes, Moraes et al. 2021). A large body of studies indicates that MDD and BD are associated with elevated peripheral levels of lipopolysaccharide (LPS), which augment inflammation and cell-mediated immunity (CMI) (Maes, Kubera et al. 2008, Maes, Twisk et al. 2012).

In comparison with simple MDD, MDD with melancholic and psychotic features and suicidal behaviors are reported to be associated with increased pro-inflammatory markers, namely APPs (e.g., haptoglobin), upregulated T cell markers, besides failure to suppress the production of IL-1β and sIL-2R by administration of dexamethasone (Maes, Bosmans et al. 1991, Maes, Lambrechts et al. 1992, Maes, Scharpé et al. 1994, Maes, Mihaylova et al. 2012, Sowa-Kućma, Styczeń et al. 2018). Hyperactive immune-inflammatory pathways may induce O&NS pathways which are accompanied by elevated reactive oxygen and nitrogen species (ROS, RNS), especially in case of lowered total antioxidant capacity levels (Maes, Galecki et al. 2011). Additionally, increased levels of myeloperoxidase (MPO), a key enzyme in the innate immune response, has been frequently reported in depression (Gałecki, Gałecka et al. 2012, Somani, Singh et al. 2022). The latter may increase reactive chlorine species (RCS, e.g. hydrochlorous acid) resulting in chlorinative stress followed by high levels of advanced oxidation protein products (AOPP) (Maes, Landucci Bonifacio et al. 2019). Moreover, high levels of oxidative mediators impact the integrity of lipids, proteins, DNA, and mitochondria (Maes, Galecki et al. 2011). Stimulated IRS and O&NS pathways explain, in part, key characteristics of affective disorders, namely the frequency of episodes (disease’s staging), the severity of illness, and suicidal behaviors, including suicidal ideation and attempts (Vasupanrajit, Jirakran et al. 2021, Maes 2022, Maes, Rachayon et al. 2022). Furthermore, the neurotoxic properties of ROS, RNS, and M1 macrophage and T helper (Th)-1 cytokines generate neuro-affective toxicity, which may explain the staging and phenome of MDD and BD (Maes 2022, Maes, Rachayon et al. 2022).

High levels of IFN-γ, IL-1β, LPS, along with ROS and RNS are implicated in induction the rate-limiting enzyme of the tryptophan catabolite (TRYCAT) pathway, namely indole 2,3-dioxygenase (IDO) enzyme (Saito, Markey et al. 1992, Anderson and Maes 2013, Reyes Ocampo, Lugo Huitrón et al. 2014, Almulla and Maes 2022). The TRYCAT pathway is the major catabolic pathway of tryptophan (TRP) which when overactive may deplete TRP thereby producing neuroactive metabolites, including kynurenine (KYN), kynurenic acid (KA), 3-hydroxykynurenine (3HK), anthranilic acid (AA), 3-hydroxyanthranilic acid (3-HA), xanthurenic acid (XA), quinolinic acid (QA), and picolinic acid (PA). The latter TRYCATs show neuroprotective as well as neurotoxic effects as shown in **Figure 1** (Maes, Leonard et al. 2011, Almulla and Maes 2022). Besides, the depletion of central and peripheral TRP levels (a precursor of serotonin) may lower serotonin levels in the CNS which have been reported in impulsive suicidal patients (Brown, Ebert et al. 1982, Maes, Leonard et al. 2011, Maes 2015, Almulla and Maes 2022). Some TRYCATs cause neuro-oxidative toxicity with oxidative cell damage and lipid peroxidation, such as 3HA, 3HK and QA (Guidetti and Schwarcz 1999, Santamaría, Galván-Arzate et al. 2001, Smith, Smith et al. 2009, Reyes Ocampo, Lugo Huitrón et al. 2014, Almulla and Maes 2022). Additionally, a substantial amount of hydrogen peroxide and superoxide anions are produced by 3HA and 3HK (Goldstein, Leopold et al. 2000).

**Figure 1:**
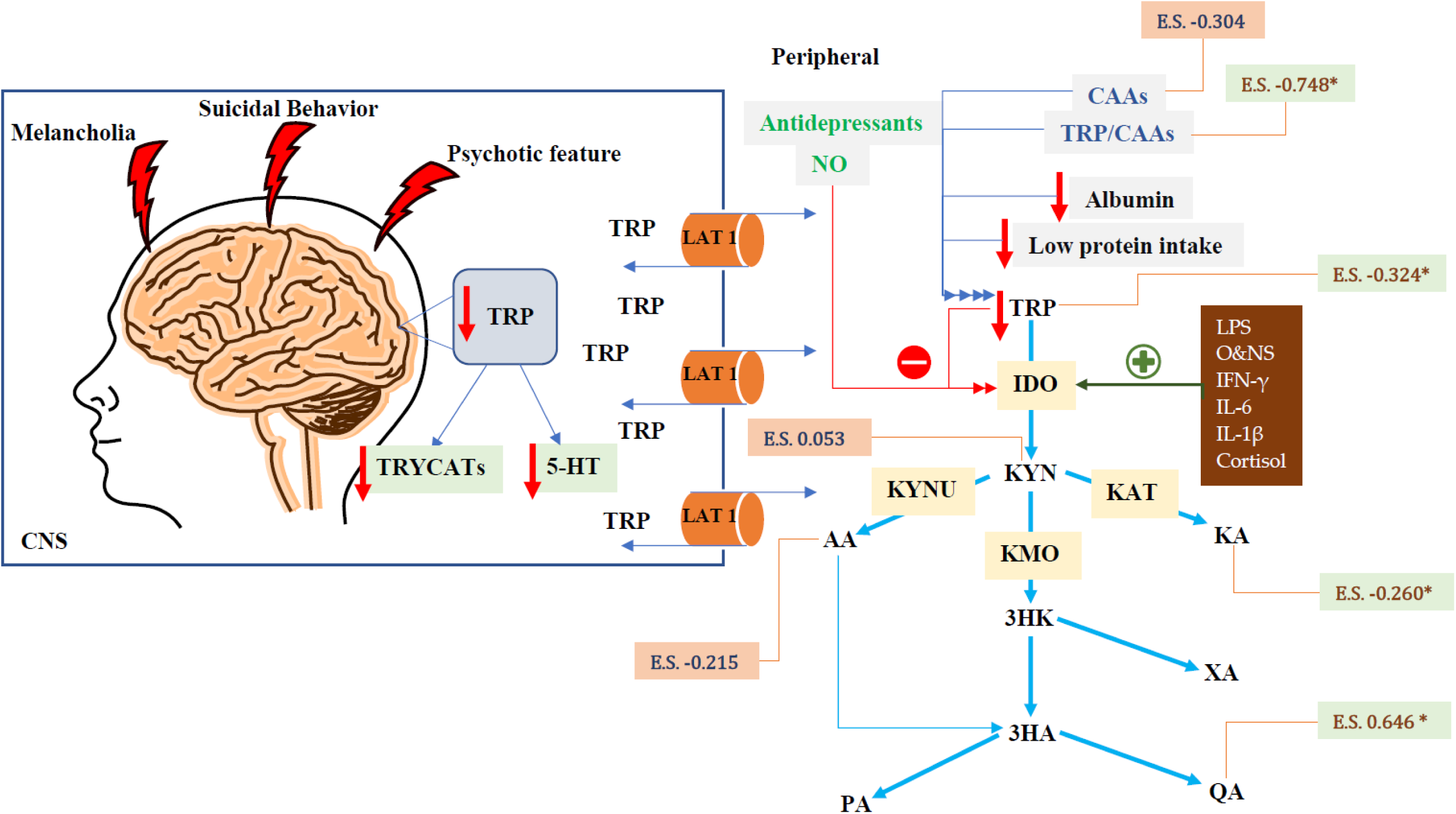
Summery of tryptophan catabolite (TRYCAT) pathway in severe affective disorders CNS: Central nervous system, TRYCATs: Tryptophan catabolites, E.S.: Effect size, TRYCAT: Tryptophan catabolite, LAT 1: Large neutral amino acid transporter 1, IFN-γ: Interferon-Gamma, IL-6: Interleukin 6, IL-1β: Interleukin-1 beta, O&NS: Oxidative and nitrosative stress, NO: Nitric Oxide, 5-HT: 5-Hydroxytryptamine, LPS: Lipopolysaccharides, CNS: Central nervous system, IDO: Indoleamine 2,3 dioxygenase, TDO: Tryptophan 2,3-dioxygenase, KAT: Kynurenine Aminotransferase, KMO: Kynurenine 3- monooxygenase, KYNU: Kynureninase, TRP: Tryptophan, KYN: Kynurenine, KA: Kynurenic Acid, 3HK: 3-Hydroxykynurenine, AA: Anthranilic Acid, XA: Xanthurenic Acid, 3HA: 3-Hydroxyanthranilic Acid, PA: Picolinic Acid, QA: Quinolinic Acid.

Furthermore, frequent agonistic effects of QA on the hippocampal N-methyl-D-Aspartate (NMDA) receptors may induce atrophy and apoptosis of the hippocampus (Maes, Leonard et al. 2011). Elevated XA levels may cause neurotoxicity by overactivation of the cationic channels that lead to intracellular hypercalcemia and, hence, accelerate damage to neural circuits in the brain along with mitochondrial dysfunction and apoptosis. These processes may substantially damage the neurons, produce poor glutamate transmission and restrict presynaptic transmission via triggering NMDA receptors (Kanchanatawan, Hemrungrojn et al. 2018, Almulla and Maes 2022). PA promotes immune-inflammatory response and reduces neuroprotection via reducing AA and KA levels which additionally have antidepressant roles (Kanchanatawan, Hemrungrojn et al. 2018, Tanaka, Bohár et al. 2020). In contrast, KYN may exert depressogenic and anxiogenic effects (Maes, Leonard et al. 2011).

We recently found, in a large-scale meta-analysis, that affective disorders, including mild to moderate severe MDD and BD, are associated with a reduction in TRP levels but without any signs of an overactivated TRYCAT pathway (Almulla, Thipakorn et al. 2022). However, in MDD patients with suicidal, melancholic, and psychotic features, central and peripheral reductions in TRP levels, activation of the TRYCAT pathway, and increased TRYCATs levels were frequently reported in previous studies, which may reveal upregulation of the IDO enzyme is the leading cause for TRP depletion and increased TRYCATs levels (Sublette, Galfalvy et al. 2011, Erhardt, Lim et al. 2013, Bay-Richter, Linderholm et al. 2015). Nonetheless, the TRP, competing amino acids (CAAs) and TRYCATs levels were not systematically reviewed in MDD/BD patients with the most severe phenotypes. Thus, in the purpose of the current study is to systematically review and meta-analyze TRP, CAAs, and the activity of TRYCAT pathway as reflected by KYN/TRP ratio (IDO enzyme index), KA/KYN (kynurenine Aminotransferase, KAT enzyme index), neurotoxicity indices and solitary levels of TRYCATs in MDD/BD patients with melancholic or psychotic features and suicidal behaviors.

## Material and methods

In the present meta-analysis, we investigated TRP, CAAs, the TRP/CAA ratio, KYN, KA, AA and QA levels, and KYN/TRP and KA/KYN ratios as indicators for IDO and KAT enzyme activities, respectively. Additionally, we also computed a neurotoxic composite, namely (KYN+3HK+3HA+QA+XA+PA). These biomarkers were examined in serum and plasma (peripherally), cerebrospinal fluid (CSF) and brain tissue (centrally) of patients with severe affective disorders who show features of melancholia, psychosis, or suicidal behavior versus healthy controls. This study followed the Preferred Reporting Items for Systematic Reviews and Meta-analyses (PRISMA) 2020 criteria (Page, McKenzie et al. 2021), the Cochrane Handbook for Systematic Reviews and Interventions guidelines (Higgins JPT 2019), and the Meta-Analyses of Observational Studies in Epidemiology (MOOSE) guidelines.

### Search strategy

ESF, Table 1 displays the keywords and MESH terms utilized to search the electronic databases, including PubMed/MEDLINE, Google Scholar, and SciFinder, for publications concerning TRP and TRYCATs in melancholia, psychotic features, and suicides of affective disorders. Moreover, to ensure the comprehensiveness of our search, we reviewed the reference lists of all eligible papers and prior meta-analyses. The current data collection processes extended from January 10 to March 31, 2022.

**Table 1.** The outcomes and number of patients with affective disorders and healthy control along with the side of standardized mean difference (SMD) and the 95% confidence intervals with respect to zero SMD.

| Outcome profiles | n<br>studies | Side of 95% confidence intervals |  |  |  | Patient<br>Cases | Control<br>Cases | Total<br>number of<br>participants |
| --- | --- | --- | --- | --- | --- | --- | --- | --- |
|  |  | < 0 | Overlap 0<br>and SMD<br>< 0 | Overlap 0<br>and SMD<br>> 0 | > 0 |  |  |  |
| TRP | 29 | 13 | 12 | 2 | 2 | 1987 | 1918 | 3905 |
| TRP/CAAs | 7 | 2 | 5 | 0 | 0 | 113 | 219 | 332 |
| CAAs | 5 | 2 | 2 | 1 | 0 | 86 | 163 | 222 |
| KYN/TRP | 17 | 1 | 5 | 7 | 4 | 1727 | 1538 | 3265 |
| KYN | 17 | 4 | 8 | 2 | 3 | 1727 | 1538 | 3265 |
| KA/KYN | 14 | 2 | 9 | 3 | 0 | 1619 | 1378 | 2997 |
| (KYN+3HK+3HA+XA+QA+PA) | 26 | 6 | 10 | 4 | 6 | 1874 | 1752 | 3626 |
| KA | 13 | 5 | 4 | 3 | 1 | 1563 | 1338 | 2901 |
| AA | 3 | 1 | 2 | 0 | 0 | 170 | 149 | 319 |
| QA | 14 | 2 | 2 | 6 | 4 | 1461 | 1131 | 2592 |

TRP: Tryptophan, KYN: Kynurenine, KA: Kynurenic acid, 3HK: 3-Hydroxykynurenine, 3HA: 3-Hydroxyanthranilic acid, XA: Xanthurenic acid, QA: Quinolinic acid, PA: Picolinic acid, AA: Anthranilic acid, CAAs: Competing amino acids (Valine + Phenylalanine + Tyrosine + Leucine + Isoleucine).

### Eligibility criteria

Articles in English that were published in peer-reviewed journals were included in our meta-analysis. Nevertheless, manuscripts in different other languages, namely Thai, French, Spanish, German, Italian, and Arabic, along with grey literature, were also selected. Other inclusion criteria were a) observational case-control and cohort studies that employed serum, plasma, CSF and brain tissues to evaluate TRP, CAAs, and/or TRYCATs, b) Diagnostic and Statistical Manual of Mental Disorders (DSM) or the International Classification of Diseases (ICD) must have been utilized to diagnose MDD and BD with either melancholia, psychotic feature and/or suicidal behavior, and c) the number of patients with melancholia, psychotic features, or suicidal behavior should be reported. Exclusion criteria were: a) genetic, animal-based and translational studies, b) research that lacked a control group, c) studies that utilized samples such as saliva, hair, whole blood, or platelet-rich plasma, and d) duplicate articles along with systematic review and meta-analyses. We emailed the authors when they did not report the mean and standard deviation (SD) or standard error (SE) of the measured biomarkers. We employed the Wan et al (Wan, Wang et al. 2014) approach to estimate the mean (SD) and the median (interquartile range or range) when we did not receive a response from authors. If graphical data were provided, the mean and SD values were extracted using the Web Plot Digitizer (https://automeris.io/WebPlotDigitizer/).

### Primary and secondary outcomes

As the primary outcomes, we examined TRP, TRP/CAAs, CAAs, KYN/TRP ratios and KYN levels in patients with affective disorders with melancholic or psychotic features, and/or suicidal behaviors versus healthy control. Secondary outcomes involved the KA/KYN ratio (KAT enzyme) and the neurotoxic composite score (KYN+3HK+3HA+QA+XA+PA) along with solitary KA, AA and QA levels.

### Screening and data extraction

Using the above inclusion criteria, the first two authors (AA and YT) performed the elementary search processes. We checked the titles and abstracts of relevant manuscripts to evaluate including them in the current meta-analysis. Once the articles passed this checking step, we downloaded the full-text articles. The first author (AA) made a Microsoft Excel file to accommodate the extracted data, mainly mean (SD) and sample size of the assessed biomarkers. Furthermore, we also recorded the medium in which the analytes were determined (serum, plasma, CSF, brain tissues), type of affective disorder and whether they showed melancholic or psychotic features, and/or suicidal behaviors, authors’ names, publication dates, location of study, and study design, as well as sex, age of the participants, psychiatric ratings scales, and the psychiatric and physical comorbidities of patients and controls. The second and third authors (YT and AV) performed double-checks for the Excel file, and they consulted the last author (MM) is case of disagreements. The last author MM, adjusted the immunological confounder scales for TRP and TRYCATs studies to evaluate the methodological quality of the included articles (Andrés-Rodríguez, Borràs et al. 2020). ESF, Table 2 shows these two rating scales, namely quality and redpoint scales which were used to examine the quality of immune-based articles on schizophrenia (Almulla, Vasupanrajit et al. 2022), Alzheimer’s disease (Almulla, Supasitthumrong et al. 2022), Coronavirus disease 2019 (Almulla, Supasitthumrong et al. 2022) and affective disorders (Almulla, Thipakorn et al. 2022). Sample size, confounder control, and the time of sampling were the main items of the quality scale, which ranged from 0 to 10, and the best quality is achieved when the score is close to 10. The redpoint scale examines the quality level of the study design in terms of major confounders, including biological and analytical bias, which can be detected by higher redpoint scale scores (ranging from 0 to 26).

**Table 2.** Results of meta-analysis performed on several outcome (TRYCATs) variables with combined different media and separately.

| Outcome feature sets | n | Groups | SMD | 95% CI | z | p | Q | df | p | I<sup>2</sup> (%) | $\tau^2$ | T |
| --- | --- | --- | --- | --- | --- | --- | --- | --- | --- | --- | --- | --- |
| TRP | 29 | Overall | -0.517 | -0.735; -0.299 | -4.650 | <0.0001 | 226.434 | 28 | <0.0001 | 87.634 | 0.282 | 0.531 |
| TRP/CAAs<sup>#</sup> | 7 | Melancholia | -0.617 | -0.957; -0.277 | -3.557 | <0.0001 | 10.435 | 6 | 0.107 | 42.500 | 0.086 | 0.293 |
| CAAs<sup>#</sup> | 5 | Overall | -0.304 | -0.674; 0.066 | -1.612 | 0.107 | 6.676 | 4 | 0.154 | 40.086 | 0.070 | 0.264 |
| KYN/TRP * | 17 | Overall | -0.032 | -0.193; 0.003 | -1.896 | 0.058 | 107.82 | 16 | <0.0001 | 85.161 | 0.170 | 0.614 |
|  | 3 | Melancholia | -0.095 | -0.193; 0.003 | -1.896 | 0.058 | 0.714 | 2 | 0.700 | 0.000 | 0.000 | 0.000 |
|  | 4 | Psychotic | 0.224 | 0.012; 0.436 | 2.068 | 0.039 | 3 | 3 | 0.392 | 0.000 | 0.000 | 0.000 |
|  | 10 | Suicidal | 0.112 | -0.119; 0.055 | -0.720 | 0.471 | 93.734 | 9 | <0.0001 | 90.398 | 0.377 | 0.614 |
| KYN | 17 | Overall | -0.114 | -0.352; 0.152 | -0.935 | 0.350 | 126.513 | 16 | <0.0001 | 87.353 | 0.203 | 0.450 |
| KA/KYN * | 14 | Overall | -0.035 | -0.117; 0.048 | -0.824 | 0.410 | 26.181 | 13 | 0.016 | 50.346 | 0.028 | 0.167 |
|  | 2 | Melancholia | 0.049 | -0.050; 0.148 | 0.969 | 0.333 | 0.402 | 1 | 0.526 | 0.000 | 0.000 | 0.000 |
|  | 5 | Psychotic | -0.201 | -0.416; 0.013 | -1.838 | 0.066 | 0.912 | 4 | 0.923 | 0.000 | 0.000 | 0.000 |
|  | 7 | Suicidal | -0.231 | -0.432; -0.030 | -2.256 | 0.024 | 13.104 | 6 | 0.041 | 54.213 | 0.039 | 0.198 |
| (KYN+3HK+3HA+XA+QA+PA) | 26 | Overall | 0.048 | -0.189; 0.284 | 0.396 | 0.692 | 209.842 | 25 | <0.0001 | 89.086 | 0.295 | 0.543 |
| KA | 13 | Overall | -0.260 | -0.487; -0.034 | -2.258 | 0.024 | 67.574 | 12 | <0.0001 | 82.242 | 0.125 | 0.354 |
| AA | 3 | Overall | -0.248 | -0.485; -0.011 | -2.055 | 0.040 | 2.115 | 2 | 0.347 | 5.432 | 0.003 | 0.051 |
| QA | 14 | Overall | 0.358 | 0.015; 0.701 | 2.044 | 0.041 | 134.272 | 13 | <0.0001 | 90.318 | 0.343 | 0.585 |

\* Significant difference between melancholia, psychotic feature and suicidal behavior

### Data analysis

ESF, Table 3 shows the PRISMA criteria which were employed in the present meta-analysis that used the CMA program V3 to analyze all of the data. The criterion to conduct a meta-analysis was that the biomarker levels should be available in at least three studies. We assumed dependency while computing the mean values of the outcomes to compare the neurotoxicity index and the KYN/TRP (an index for IDO enzyme activity) and KA/KYN (an index for KAT enzyme activity) ratios in depressed patients versus healthy controls (Almulla, Supasitthumrong et al. 2022, Almulla, Vasupanrajit et al. 2022). We evaluate the following ratios by specifying the effect size direction a) KYN/TRP with KYN in positive direction and TRP in negative direction, b) KA/KYN with KA in positive and KYN in negative direction; c) TRP/CAAs ratio: TRP in positive and CAA in negative direction. We employed the random-effects model with restricted maximum likelihood to extract the effect size and report the standardized mean difference (SMD) with 95 percent confidence intervals (95% CI) as an indicator for the effect size with a two-tailed p-value less than 0.05 to describe the statistical significance. According to the values of SMD, namely 0.2, 0.5, and 0.8, the effect sizes were considered small, medium, and large, respectively (Cohen 1988). We used tau squared statistics to delineate heterogeneity in the data but also assessed the Q and I^2^ metrics (Almulla, Supasitthumrong et al. 2022, Almulla, Vasupanrajit et al. 2022, Vasupanrajit, Jirakran et al. 2022). We also performed meta-regression analyses to detect the sources of heterogeneity. Subgroup analysis was utilized to find the variations in TRP and TRYCATs among patients with melancholia, psychotic features, and suicidal behavior and central nervous system (CNS, brain tissues + CSF), serum and plasma, while selecting each of the latter groups as a unit of analysis. Since we did not find any significant difference between CSF and brain tissues, we combined the results from CSF and brain tissues into one group, called CNS. The effect sizes obtained from melancholic, psychotic, and suicidal patients were pooled in the absence of any significant difference between the above groups. Nevertheless, if there were significant intergroup differences, we report the effect sizes separately in the various subgroups. The strength of the effect sizes was examined by carrying out sensitivity analysis utilizing a leave-one-out approach. The fail-safe N technique along with one-tailed p-values for Kendall tau with continuity correction and Egger’s regression intercept were computed to investigate possible publication bias. The adjusted effect sizes were computed after imputing missing studies by the trim-and-fill method when the Egger’s test showed significant asymmetry. Funnel plots are generated and show study precision plotted versus SMD (with both observed and imputed values).

**Table 3.** Results on publication bias.

| Outcome feature sets | Fail safe n | Z Kendall's $\tau$ | p | Egger's t test (df) | p | Missing studies (side) | After Adjusting | |
| --- | --- | --- | --- | --- | --- | --- | --- | --- |
|  |  |  |  |  |  |  | SMD | 95%CI |
| TRP | -10.30 | 1.819 | 0.034 | 3.063 (27) | 0.002 | 5 (Right) | -0.324 | -0.548; -0.101 |
| TRP/CAAs | -4.785 | 0.300 | 0.381 | 1.142 (5) | 0.152 | 2 (Left) | -0.748 | -1.069; -0.427 |
| KYN/TRP (Psychotic) | 1.827 | 0.339 | 0.367 | 0.764 (2) | 0.262 | - | - | - |
| KA/KYN (Suicidal) | -3.342 | 0.600 | 0.274 | 0.663 (5) | 0.268 | 2 (Left) | -0.337 | -0.541; -0.133 |
| (KYN+3HK+3HA+XA+QA+PA) | -0.094 | 0.705 | 0.240 | 0.892 (24) | 0.190 | 3 (Right) | 0.210 | -0.046; 0.467 |
| KA | -5.072 | 0.549 | 0.291 | 0.692 (11) | 0.251 | - | - | - |
| AA | -2.199 | 0.000 | 0.500 | 0.422 (1) | 0.372 | 1 (Right) | -0.215 | -0.430; 0.00006 |
| QA | 3.846 | 0.985 | 0.162 | 1.618 (12) | 0.065 | 3 (Right) | 0.646 | 0.228; 1.065 |

TRP: Tryptophan, KYN: Kynurenine, KA: Kynurenic acid, 3HK: 3-Hydroxykynurenine, 3HA: 3-Hydroxyanthranilic acid, XA:
Xanthurenic acid, QA: Quinolinic acid, PA: Picolinic acid, AA: Anthranilic acid, CAAs: Competing amino acids (Valine +
Phenylalanine + Tyrosine + Leucine + Isoleucine).

## Results

### Search results

The number of included and excluded studies and the final search outcome are displayed in the PRISMA flow chart **Figure 2**. We employed MESH terms and keywords (all shown in ESF, Table 1) to perform the initial search process, including inspection of 10861 articles. Based on our exclusion criteria 35 studies were selected as eligible in our systematic review and meta-analysis (Cowen, Parry-Billings et al. 1989, Anderson, Parry-Billings et al. 1990, Maes, Jacobs et al. 1990, Price, Charney et al. 1991, Quintana 1992, Maes, Meltzer et al. 1993, Møller 1993, Maes, De Backer et al. 1995, Maes, Wauters et al. 1996, Song, Lin et al. 1998, Hoekstra, Fekkes et al. 2006, Miller, Llenos et al. 2006, Myint, Kim et al. 2007, Myint, Kim et al. 2007, Gabbay, Klein et al. 2010, Steiner, Walter et al. 2011, Sublette, Galfalvy et al. 2011, Erhardt, Lim et al. 2013, Bay-Richter, Linderholm et al. 2015, Bradley, Case et al. 2015, Busse, Busse et al. 2015, Dahl, Andreassen et al. 2015, Savitz, Drevets et al. 2015, Brundin, Sellgren et al. 2016, Clark, Pocivavsek et al. 2016, Zhou, Zheng et al. 2018, Aarsland, Leskauskaite et al. 2019, Pompili, Lionetto et al. 2019, Sellgren, Gracias et al. 2019, Achtyes, Keaton et al. 2020, Ryan, Allers et al. 2020, van den Ameele, van Nuijs et al. 2020, Milaneschi, Allers et al. 2021, Trepci, Sellgren et al. 2021).

**Figure 2:**
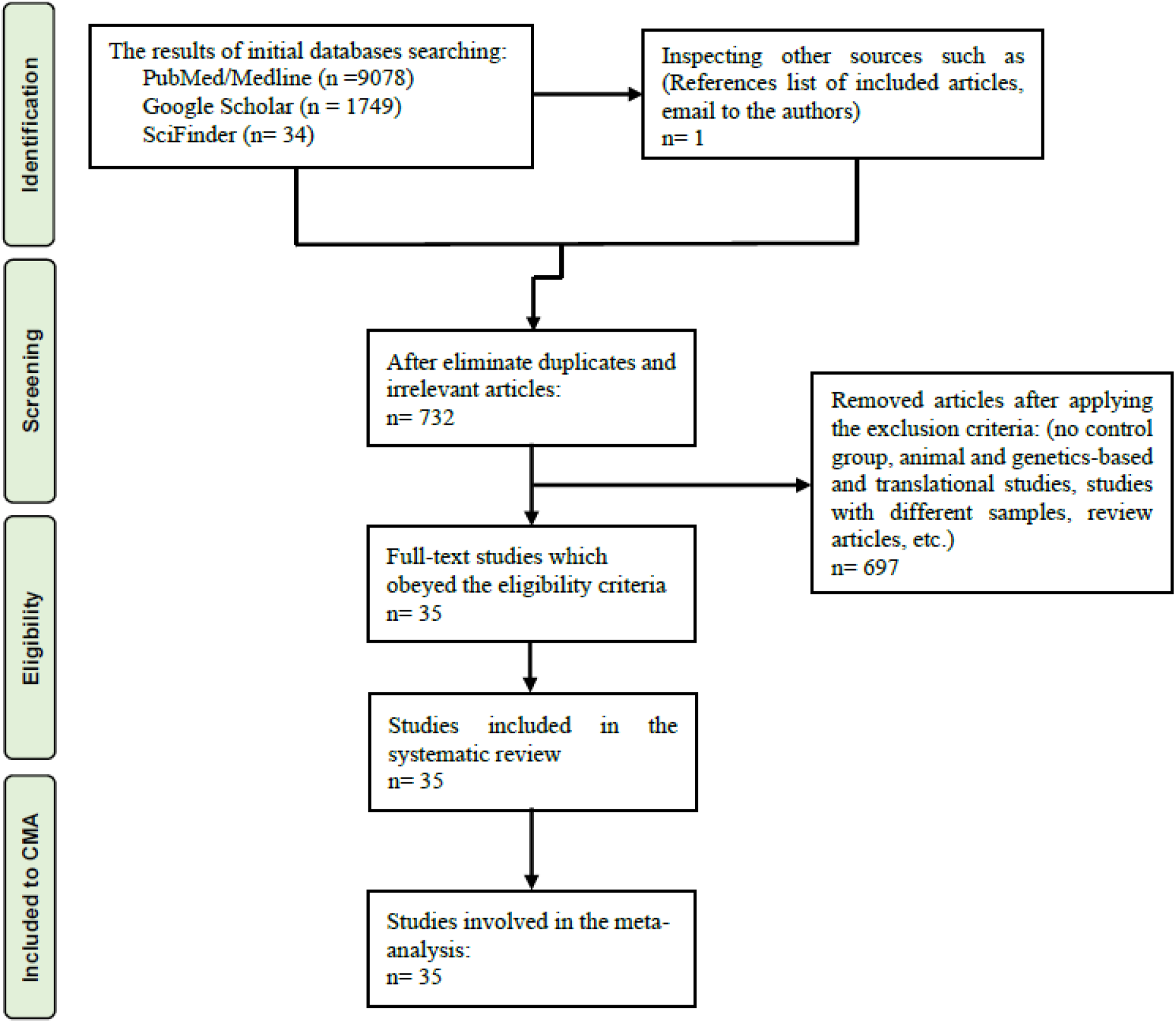
The PRISMA flow chart.

One of the included studies reported two separate cohorts of patients namely those with melancholia and psychotic features. Therefore, we entered it two times and thus we pooled the overall effect sizes from 36 studies in the present systematic review and meta-analysis (8 CNS, 21 plasma, and 7 serum). We included 11 studies on depression with melancholia (9 plasma and 2 serum), 8 studies of depression with psychotic features (1 CNS, 5 plasma and 2 serum) and 17 studies with suicidal behavior (7 CNS, 7 plasma and 3 serum). The current meta-analysis included 4,647 participants distributed as 2,332 patients and 2,315 healthy controls. The mean ages of the individuals in the studies extended from 30 to 59 years.

ESF, Table 4 shows that USA, Belgium, and Sweden contributed most to the total number of studies (8, 6 and 5 studies), respectively. Norway, the United Kingdom, Germany, Netherlands, and South Korea contributed with 2 studies and Italy, Spain, Ireland, China and Tunisia contributed each one study. High-performance liquid chromatography (HPLC) has been used in 14 studies and was, therefore, the most commonly technique employed to assess TRP and TRYCATs (see ESF, table 4). This table also shows the quality (median=5.62, min=2.75, max=7.75) and redpoint (median=13.75, min=9.5, max=18.5) scores.

### Primary outcome variables

#### TRP, CAAs levels and the TRP/CAAs ratio

**Table 1** shows that the effect size of the TRP level was pooled from 29 studies. The CI was completely on the negative side of zero in 13 studies, whereas only 2 studies showed that the CIs were on the positive side of zero. Furthermore, 14 studies intersected with zero with a negative SMD in 12 and a positive SMD in 2 studies. TRP levels were significantly decreased with a moderate effect size (SMD= -0.517) in patients compared to healthy controls. **Figure 3** shows the forest plot of the TRP results. Publication bias analysis revealed 5 missing studies to the right side of the funnel plot and imputing these studies resulted in a lowered effect size although it remained significant.

**Figure 3:**
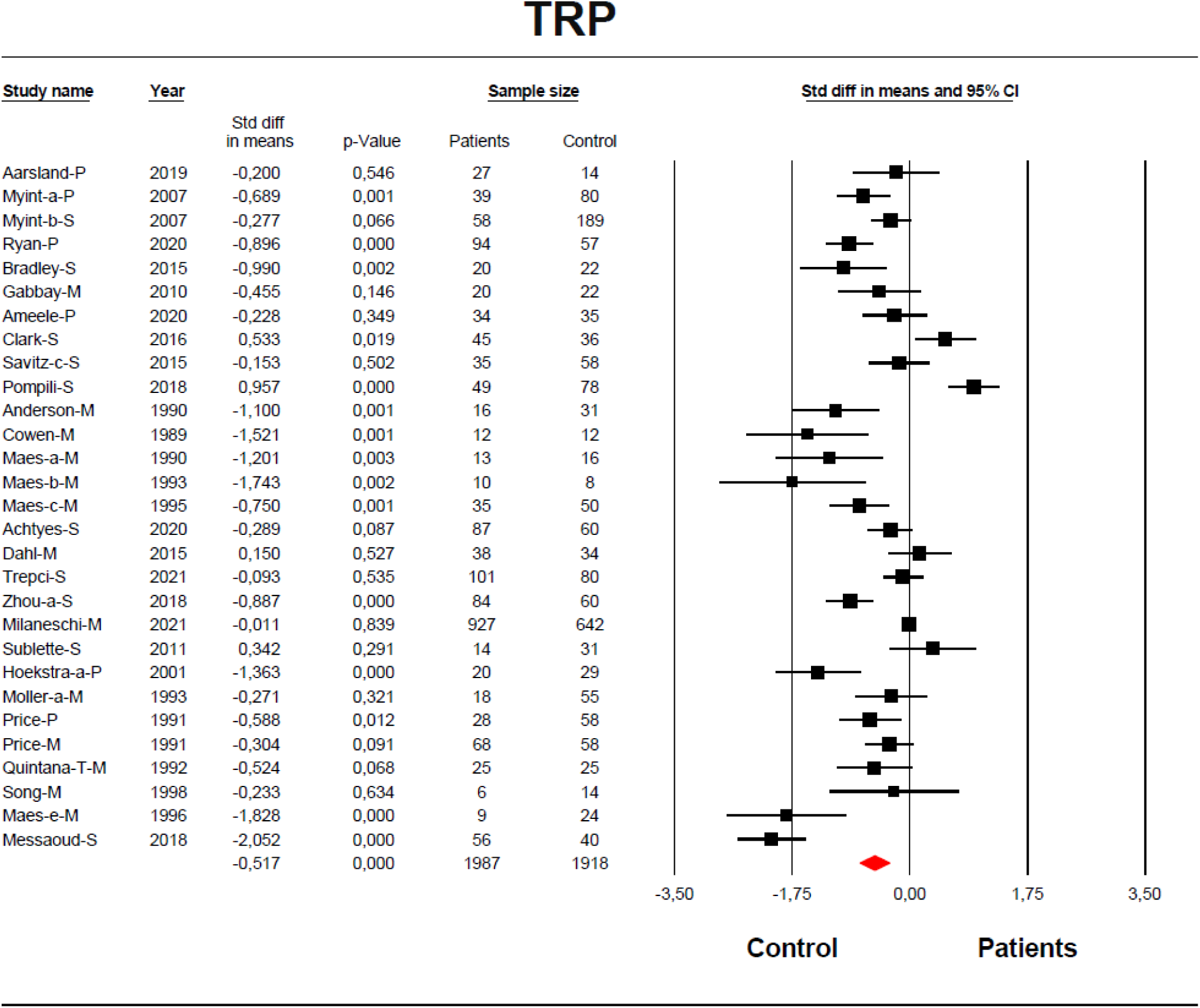
The forest plot of tryptophan (TRP) between severe affective disorder patients and healthy control.

CAAs results were obtained from 5 studies which involved only MDD with melancholia. Table 1 shows that the CIs of 2 studies fell entirely on the negative side of zero and 3 studies intersected with zero, 2 with negative and 1 with positive SMD values. ESF, Figure 1 shows that CAAs was not significantly different between patients and controls.

The effect size of TRP/CAAs ratio was extracted from 7 studies performed in MDD with melancholia features. The 95% CI was entirely on the negative size of zero in 2 studies and 5 studies showed an overlap with zero. Melancholic MDD patients showed a significant reduction in TRP/CAAs with moderate effect size (SMD= -0.617). There were 2 missing studies on the left side of the funnel plot and after adjusting the effect size for these missing studies the SMD value was -0.748.

### The KYN/TRP ratio and KYN levels

**Table 1** and **Figure 4** revealed no significant differences in the KYN/TRP ratio between patients and controls. Due to the high heterogeneity, we performed group analysis which showed significant differences between melancholic, suicidal and psychotic patients and a significant increase with a small effect size was established in psychotic depression. Z Kendall’s and Egger’s test showed no signs of publication bias.

**Figure 4:**
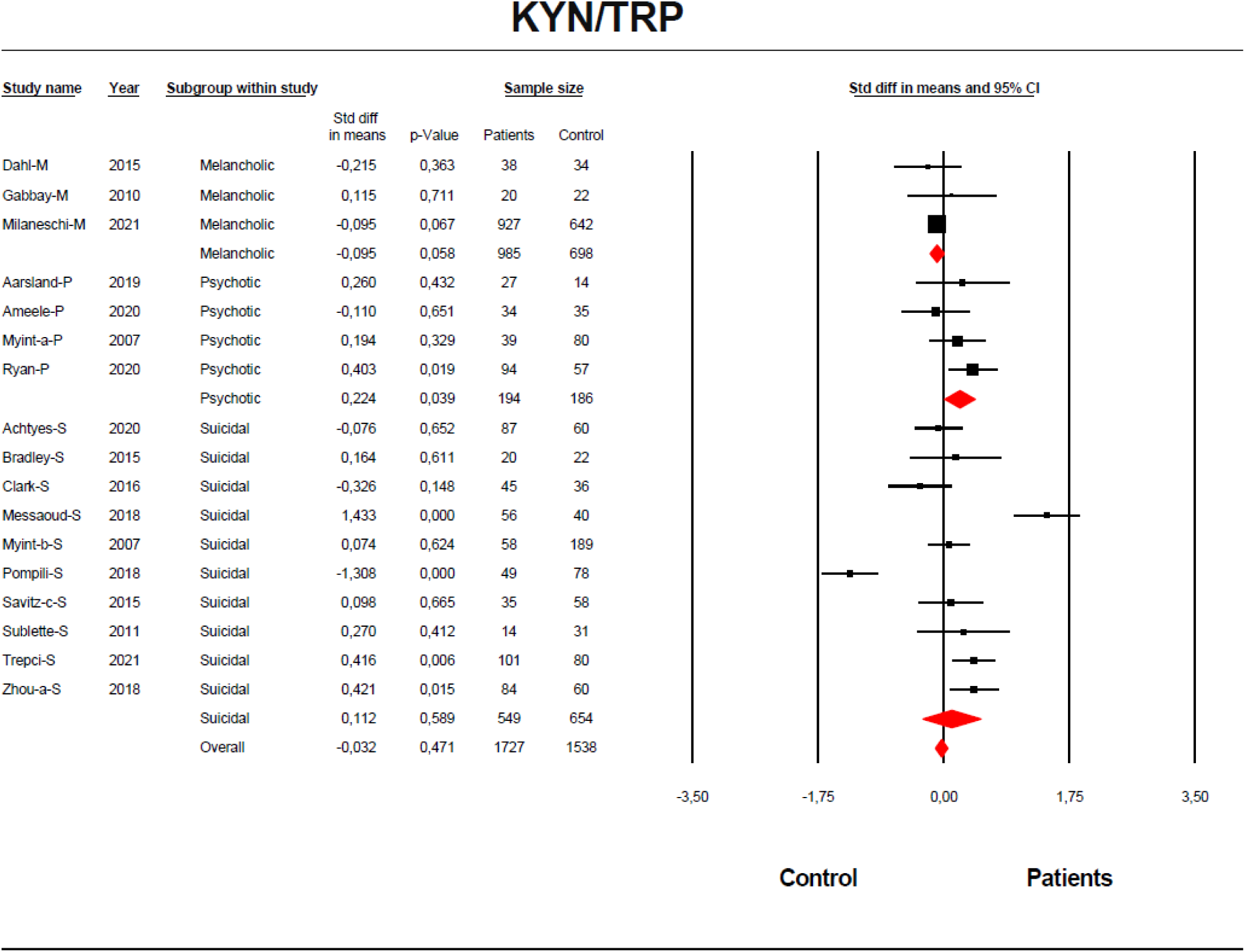
The forest plot of kynurenine (KYN)/tryptophan (TRP) ratio between severe affective disorder patients and healthy control.

**Table 1** and **2** and ESF, Figure 3 show no significant difference in KYN levels between patients and controls. Group analysis displayed a significant difference (p=0.041) between CNS, serum and plasma KYN levels which were significantly decreased in plasma. **Table 3** revealed 3 missing studies on the right side of funnel plot and imputing these missing data yielded a non-significant effect size.

### Secondary outcome variables

#### Neurotoxicity composite (KYN+3HK+3HA+XA+QA+PA) and KA/KYN ratio

Table 1 and ESF, figure 4 show no significant differences in this neurotoxicity composite between patients and controls. We obtained the effect size of KA/KYN ratio from 14 studies and Table 2 and ESF, Figure 5 indicate no overall difference between patients and controls. However, group analysis showed a significant difference between melancholic, psychotic and suicidal patients, while only the latter displayed a significant decrease with a small effect size in the KA/KYN ratio compared to controls. Table 3 shows that there were 2 missing studies on the left side of the funnel plot of the KA/KYN ratio in suicidal patients and the adjusted estimate value was more decreased after imputing these missing studies.

### Solitary levels of KA, AA and QA

Table 2 and ESF, Figure 6 show that KA levels in patients were significantly lower with a small effect size in patients as compared with controls. **Table 1** and ESF, Figure 7 show that the effect size of AA was obtained from three studies and that AA is significantly decreased with small effect size in patients. Table 3 shows one missing study to the right side of the funnel plot and after imputing this missing study showed that the results were no longer significant. **Table 1 and 2** and ESF, Figure 8 show that the QA levels were significantly increased with small effect size in patients versus controls. Publication bias analysis showed that there were 3 missing studies on the right of the funnel plot and after imputing these studies, the effect size increased to 0.646.

### Meta-regression analyses

In order to examine factors which could explain heterogeneity, we carried out meta-regression analyses (ESF, table 5). Plasma was the most important confounder increasing heterogeneity with significant effects on TRP, KYN, KA, QA and (KYN+3HK+3HA+XA+QA+PA). Moreover, male/female gender, absence of electroconvulsive therapy (ECT), medications and latitude also impact heterogeneity.

## Discussion

### Availability of TRP to the brain

The first major finding in the current study is that TRP levels and the TRP/CAA ratio are significantly decreased in patients with MDD/BD with melancholia and psychotic features, and suicidal behaviors as compared with healthy controls and that there was no significant difference between the latter phenotypes in TRP and TRP/CAA levels. The current results align with findings of a recent study conducted on MDD and BD patients with mild to moderate forms of depression (Almulla, Thipakorn et al. 2022). Moreover, previous studies revealed a significant TRP reduction in affective disorders with melancholic, psychotic and suicidal features (Maes, Wauters et al. 1996, Hoekstra, Fekkes et al. 2006, Gabbay, Klein et al. 2010, Bradley, Case et al. 2015). We also found that melancholic MDD patients have normal levels of CAAs and a decreased TRP/CAAs ratio, which is therefore solely determined by diminished TRP levels. These findings are consistent with our recent findings in mild to moderate MDD/BD (Almulla, Thipakorn et al. 2022). We could not examine the CAA levels and TRP/CAA ratio in MDD/BD patients with psychotic features and/or suicidal behaviors because there were not sufficient studies. Thus, the current meta-analysis confirms previous studies about MDD patients with melancholia (Maes, Jacobs et al. 1990, Maes, Meltzer et al. 1993, Maes, De Backer et al. 1995, Maes, Wauters et al. 1996).

It is important to measure the TRP/CAA ratio because both peripheral TRP (whether total or free) and CAA (tyrosine, valine, phenylalanine, leucine and isoleucine) levels determine at least in part brain TRP concentration (Yuwiler, Oldendorf et al. 1977, Pardridge 1979). Indeed, specific receptors in the blood-brain barrier (BBB), named the large amino acid transporter 1 (LAT 1), are responsible for delivering TRP to the brain from the peripheral blood and the above CAAs compete with TRP to the cross BBB (Almulla, Vasupanrajit et al. 2022). In this respect, Moller (Møller 1985) reported that a decreased TRP/CAAs ratio predicted a successful response to the selective serotonin reuptake inhibitors (SSRIs).

### KYN levels and IDO enzyme

The second major finding of the present study is that patients with severe affective disorders showed unchanged KYN levels compared with healthy controls. However, these results showed high heterogeneity, and, hence, we performed group analysis which revealed that only plasma KYN level was significantly decreased which is consistent with previous results in mild to moderate MDD/ BD (Almulla, Thipakorn et al. 2022, Almulla, Vasupanrajit et al. 2022). We also found that the KYN/TRP ratio was unaltered in all patients combined although patients with psychotic features showed a trend towards significant increases in the KYN/TRP ratio. Nonetheless, since KYN is not significantly increased, such changes do not result from IDO activation.

While some prior studies are in line with our findings and showed neither significant changes in KYN/TRP ratio nor KYN level in suicidal patients (Savitz, Drevets et al. 2015, Trepci, Sellgren et al. 2021), Messaoud et al. reported a significant plasma elevation of KYN level and KYN/TRP ratio in suicidal MDD patients (Messaoud, Mensi et al. 2019). However, another study showed a high serum KYN/TRP ratio without changes in KYN level (Zhou, Zheng et al. 2018). The KYN/TRP ratio was significantly increased in MDD patients with psychotic features, while the KYN level decreased significantly (Ryan, Allers et al. 2020). Other studies that included melancholic patients reported no significant changes in plasma KYN levels and the KYN/TRP ratio (Dahl, Andreassen et al. 2015, Milaneschi, Allers et al. 2021).

The current meta-analysis indicates that there is no overactivation of the TRYCAT pathway in severe forms of affective disorders, which are, nevertheless, accompanied by a mild chronic immune-inflammatory response. Similar findings were recently reported in Alzheimer’s disease, another disorder accompanied by a mild chronic inflammatory process (Almulla, Supasitthumrong et al. 2022, Almulla, Thipakorn et al. 2022). In contrast, the TRYCAT pathway is more pronounced in conditions characterized by severe acute inflammatory conditions such as COVID-19 infection (Almulla, Supasitthumrong et al. 2022) and treatment with IFN-α (Bonaccorso, Marino et al. 2002). In those conditions, cytokine induced TRP depletion (also known as TRP starvation) is a key element in the innate immune response that impedes intruding pathogens and results in anti-inflammatory effects (Maes, Leonard et al. 2011, Almulla and Maes 2022).

Nevertheless, several factors influence the activity of the IDO enzyme and probably lead to inhibition of the TRYCAT pathway. First, lowered TRP levels may drive the self-regulation of the IDO enzyme which is accompanied by an inactive ferric IDO form and autoxidation (Booth, Basran et al. 2015, Nelp Micah, Kates Patrick et al. 2018). Second, elevated nitric oxide levels are reported in severe MDD and BD and may inhibit the IDO enzyme (Kim, Paik et al. 2006, Lee, Lee et al. 2006, Maes, Simeonova et al. 2019). Third, cellular IDO is probably inhibited at the post-translational level by high hydrogen peroxides concentrations (Freewan, Rees et al. 2013) which is another hallmark of depression (Leonard and Maes 2012). Moreover, other substantial factors which influence the TRYCAT pathway are deficiencies of riboflavin (vitamin B2), a coenzyme of kynurenine 3-monooxygenase (KMO), and pyridoxal 5′-phosphate (PLP, vitamin B6) which is the coenzyme for KAT and kynureninase (KYNU) (Ryan, Allers et al. 2020). Both vitamins are repeatedly reported to be decreased in depression (Naghashpour, Amani et al. 2011, Mikkelsen, Stojanovska et al. 2017).

Since IDO is not activated in severe depression, other factors should explain the lower TRP availability to the brain. First, depression is accompanied by lowered levels of albumin, a negative APP, whilst a large part of the TRP pool is bound to albumin in the peripheral circulation (Mc and Oncley 1958, Fernstrom, Larin et al. 1973). As such, lowered albumin will decrease total TRP levels, thereby lowering brain TRP concentrations and maybe impacting serotonin synthesis in the brain (Maes, Vandewoude et al. 1991, Maes, Wauters et al. 1996, Maes, Smith et al. 1997). In this respect, lowered albumin showed a negative correlation with depressive symptoms in patients with suicidal attempts (Ambrus and Westling 2019). Second, platelet uptake of TRP may be increased in MDD (Morel-Kopp, McLean et al. 2009, Moreno, Gaspar et al. 2013). Third, increased free fatty acids, partially mediated by insulin, may affect TRP levels in MDD (Almulla, Vasupanrajit et al. 2022). All the above-mentioned causes are probably responsible for reduced levels of TRP in severe affective disorders in the absence of upregulated TRYCAT pathway.

It should be stressed that patients with severe forms of affective disorders are probably treated with many antidepressants, mood stabilizers and antipsychotics, which may have substantial effects on IDO activity and TRYCATs levels (Ara and Bano 2012, Qiu, Yang et al. 2014). Moreover, some antidepressants have anti-inflammatory properties by a) inhibiting overactivated cell-mediated immunity and decreasing IFN-γ levels (a potent stimulator of IDO enzyme), therefore impeding stimulation of the TRYCAT pathway (Maes 2011), and b) reducing various acute-phase proteins, such as haptoglobin, fibrinogen, C3, C4, and α-antitrypsin (Maes, Delange et al. 1997). Furthermore, animal-based studies reported that valproate and citalopram negatively regulate the IDO and tryptophan 2,3-dioxygenase (TDO) enzymes and reserve TRP for the 5-HT pathway (Ara and Bano 2012, Qiu, Yang et al. 2014). Therefore, in severe affective disorders, many factors control the activity of the TRYCAT pathway.

### Neurotoxic indexes and TRYCATs

The third major finding of the current study is that the neurotoxicity index (including the composite score KYN+3HK+3HA+XA+QA+PA) is unaltered in patients with severe affective disorders. In addition, the KA/KYN ratio (another neurotoxicity index) was significantly decreased in patients with suicidal behavior and psychotic features but not in those with melancholia. The alteration in the KA/KYN ratio is probably due to decreased KA levels since we did not find any abnormality in KYN levels. We recently found that mild to moderate MDD/BD patients showed unchanged neurotoxicity composite scores and an increased KA/KYN ratio (Almulla, Thipakorn et al. 2022). In line with the current study, previous studies showed a reduced KA/KYN ratio in suicidal depressed patients (Zhou, Zheng et al. 2018).

Nevertheless, severe affective disorder was accompanied by a significantly increased level of QA, a neurotoxic TRYCAT. Previous studies concerning QA were often inconsistent. For example, in the CSF of suicidal MDD patients, QA levels were elevated (Erhardt, Lim et al. 2013, Bay-Richter, Linderholm et al. 2015), while postmortem studies of suicide patients showed that in the CA1 and CA2/3 areas of the hippocampus, QA levels were either significantly decreased or unchanged (Busse, Busse et al. 2015). In contrast, QA levels were increased in subgenual anterior cingulate cortex (sACC), and anterior midcingulate cortex (aMCC) areas of the brain of depressed patients with suicidal behaviors (Steiner, Walter et al. 2011). Moreover, in the plasma of suicidal MDD patients, QA levels were either significantly decreased (Achtyes, Keaton et al. 2020) or unaltered (Brundin, Sellgren et al. 2016). No significant changes in QA levels were found in depressed patients with psychotic features or melancholia (Dahl, Andreassen et al. 2015, Aarsland, Leskauskaite et al. 2019). In the current study, we did not find any difference between peripheral and central QA levels or between patients with melancholia, psychotic features, and suicidal behavior, but, overall, there was a significant increase.

### Neuroprotective TRYCATs

The fourth major finding of this study is that severe forms of affective disorders are linked to lower KA levels without a change in AA levels (after bias correction), even though the effect size was only computed in 3 studies. Recently, we found in mild to moderate MDD and BD patients that there was a peripheral reduction in KA and no changes in central levels (Almulla, Thipakorn et al. 2022). In this regard, we did not find any significant difference among central and peripheral levels in the present study. Some previous papers indicated significantly lower central and peripheral KA levels (Bay-Richter, Linderholm et al. 2015, Aarsland, Leskauskaite et al. 2019). However, other studies showed no aberration in KA levels, whether centrally or peripherally, in suicidal and melancholic MDD patients (Erhardt, Lim et al. 2013, Dahl, Andreassen et al. 2015). The main cause of lower KA is probably a decrease in KYN, the substrate of the KAT enzyme, although dietary factors cannot be excluded (Tomaszewska, Muszyński et al. 2019). Recently, Steiner et al found a strong negative correlation between AA and severity of depression scores in unmedicated depressed females implying that the severity of depression is associated with lowered AA levels (Steiner, Dobrowolny et al. 2021).

All in all, it appears that MDD/BD may be accompanied by lowered neuroprotection and consequent increased neurotoxicity. First, our findings indicating decreased KA but increased QA in severe affective patients indicate QA-based neurotoxicity (Almulla and Maes 2022). Second, lowered levels of TRP (in itself an antioxidant) may lead to decreased antioxidant metabolites, namely serotonin, melatonin, 3HK and XA (Reiter, Tan et al. 1999, Xu, Liu et al. 2018). Moreover, serotonin enhances proper neuroplasticity and maintains healthy neurons (Croonenberghs, Verkerk et al. 2005, Rădulescu, Drăgoi et al. 2021). Third, KA has a neuroprotective role by antagonizing the action of QA inhibiting excitatory receptors namely NMDA, α-amino-3-hydroxy-5- methyl-4-isoxazolepropionic acid (AMPA) and kainate glutamate ionotropic receptors, in addition, to impede alpha 7 nicotinic acetylcholine receptor (α7nAChr) and thereby decrease glutamate release (Morris, Carvalho et al. 2016, Almulla and Maes 2022). Fourth, inhibition of the TRYCAT pathway results in decreased KYN, KA and XA levels, which is associated with an indirect increase in neurotoxicity since these metabolites exert an anti-inflammatory role by diminishing IFN-γ/IL-10 ratio (Maes, Mihaylova et al. 2007). In addition, KA, 3HK, 3HA, and XA display antioxidant properties (Goda, Hamane et al. 1999, Maes, Leonard et al. 2011).

## Limitations

Some limitations should be noticed while interpreting the current findings. The lack of information concerning treatment histories restricted our potential to examine the impact of drugs on TRP, CAAs and TRYCATs levels. We could not examine the status of TRP and TRYCATs in the central nervous system because only a limited number of studies on CSF and brain were available. Furthermore, no studies assessed CAAs in MDD with psychotic features or suicidal behaviors, and, therefore, we could not examine CAA levels and the TRP/CAAs ratio in these subgroups.

## Conclusions

Figure 1 shows the summary of our findings. Severe affective disorders are accompanied by a decreased availability of TRP to the brain, whilst the TRYCAT pathway is not upregulated, probably due to the multiple treatments administered to those patients. However, there was a significant increase in neurotoxic QA and a significant decrease in neuroprotective KA, indicating increased neurotoxicity.

## Ethical approval and consent to participate

Not applicable.

## Consent for publication

Not applicable.

## Availability of data and materials

The dataset generated during and/or analyzed during the current study will be available from the corresponding author (MM) upon reasonable request and once the dataset has been fully exploited by the authors.

## Author’s contributions

The design of the study was made by AA and MM. AA and YT searched and collected the relevant data. AA and MM conducted the statistical analysis. All authors contributed to the writing of this study.

## Acknowledgments

Not applicable.

