## supplementary file for "The tryptophan catabolite or kynurenine pathway in a major depressive episode with melancholia, psychotic features and suicidal behaviors; a systematic review and meta-analysis"

**SHORT TITLE: Kynurenine pathway in affective disorders**

Abbas F. Almulla, Ph.D.[a,b] , Yanin Thipakorn, M.D., Ph.D.[a], Asara Vasupanrajit, MS.c[a], , Chavit Tunvirachaisakul, M.D., Ph.D.[a], Gregory Oxenkrug, M.D., Ph.D.[c], Hussein K. Al-Hakeim, Ph.D.[d], Michael Maes, M.D., Ph.D.[a,e,f]

[a] Department of Psychiatry, Faculty of Medicine, Chulalongkorn University, Bangkok, Thailand.

[b] Medical Laboratory Technology Department, College of Medical Technology, The Islamic University, Najaf, Iraq.

[c] Department of Psychiatry, Tufts University School of Medicine and Tufts Medical Center, Boston, MA 02111, USA.

[d] Department of Chemistry, College of Science, University of Kufa, Kufa, Iraq.

[e] Department of Psychiatry, Medical University of Plovdiv, Plovdiv, Bulgaria.

[f] Deakin University, IMPACT - the Institute for Mental and Physical Health and Clinical Translation, School of Medicine, Barwon Health, Geelong, Australia.

**Corresponding author:**

Prof. Dr Michael Maes, M.D., Ph.D.

Department of Psychiatry

Faculty of Medicine, Chulalongkorn University

Bangkok, 10330

Thailand

E-mail addresses:

**ESF, Table 1.** Search sentences and terms were used in each database

| Database Name | Search Sentence | No. of Articles |
|---|---|---|
| **PubMed/Medline** | ((((((((((TRYCATs and Suicide)* OR (kynurenine pathway and Suicide)) OR (Tryptophan Catabolites and Suicide)) OR (L-TRP and Suicide)) OR (L-KYN and Suicide)) OR (KYNA and Suicide)) OR (ANA and Suicide)) OR (3-HK and Suicide)) OR (XA and Suicide)) OR (3-HA and Suicide)) OR (QA and Suicide)) OR (PA and Suicide) | **7714** |
| | (((((((((Melancholia and TRYCATs)* OR (Melancholia and Tryptophan)) OR (melancholia and tryptophan cata)) OR (Melancholia and TRYCATs)) OR (Melancholia and TRP)) OR (Melancholia and KYN)) OR (Melancholia and kynurenine)) OR (Melancholia and Kynurenine pathway)) OR (Melancholia and IDO)) OR (Melancholia and TDO) | **1283** |
| | (((((((((Psychotic depression and TRYCATs)* OR (Psychotic depression and Tryptophan)) OR (Psychotic depression and tryptophan cata)) OR (Psychotic depression and TRYCATs)) OR (Psychotic depression and TRP)) OR (Psychotic depression and KYN)) OR (Psychotic depression and kynurenine)) OR (Psychotic depression and IDO)) OR (Psychotic depression and TDO) | **81** |
| **Google Scholar** | (((((Melancholia* and IDO activation) OR (TDO activation)) OR (Kynurenine pathway)) OR (KMO activation)) OR (Tryptophan degradation)) AND (Decreased Tryptophan) | **499** |
| | (((((Psychotic depression * and IDO activation) OR (TDO activation)) OR (Kynurenine pathway)) OR (KMO activation)) OR (Tryptophan degradation)) AND (Decreased Tryptophan) | **1250** |
| **SciFinder** | Suicide and kynurenine pathway, Suicide and tryptophan catabolism pathway, Suicide and TRYCATs, Suicide and kynurenine, Suicide and tryptophan, Suicide kynurenic acid, Suicide and kynurenine pathway | **9** |

| | | |
|---|---|---|
| | Melancholia and kynurenine pathway, Melancholia and tryptophan catabolism pathway, Melancholia and TRYCATs, Melancholia and kynurenine, Melancholia and tryptophan, Melancholia and kynurenic acid, Psychotic depression and kynurenine pathway | **25** |

**ESF, Table 2.** Immune cofounder's scale (ICS); adapted from Andrés-Rodríguez, et al., 2019

| **Methodological quality of the study** | |
|---|---|
| **1** | Study sample ≥ 128 participants including patients and controls (1= Yes, 0 = No) |
| **2** | Did the study control the results for potential confounders (e.g., age, BMI, gender, race)? (1= Yes, 0 = No) |
| **3** | Were participants with affective disorders and controls age- and-gender-matched or was there a statistical control? (1= Yes, 0 = No) |
| **4** | Was the time of sample collection specified (e.g., morning vs. evening)? (1= Yes, 0 = No) |
| **5** | Were participants with affective disorders free of immunomodulatory drugs including anti-cytokines, glucocorticoids, immunoglobulins, and immunosuppressants, or was there a medication washout period, or was drug intake statistically controlled for? (1= Yes, 0 = No) |
| **6** | Were participants with affective disorders free of antidepressants and mood stabilizers or were the data statistically controlled for? (1= Yes, 0 = No) |
| **7** | Reporting either the manufacturer of the test or detection limit and coefficients of variation (1= Yes, 0 = No) |
| **8** | Reporting how data under detection limit were handled (1 = Yes, 0 = No) |
| **9** | Reporting % of the sample under detection limit (1=Yes, 0= No) |
| **10** | Reporting blood fraction (serum, plasma, culture supernatant or whole blood) (1= Yes, 0 = No) |
| | **Total quality score (10 points)** |
| **Biomarker confounders red points** | |

| *The red points should not be given if the item is statistically controlled for* | |
|---|---|
| **1** | 3 red points for comorbid illnesses such as autoimmune disorders & other immune disorders including rheumatoid arthritis, psoriasis, inflammatory bowel disease, chronic obstructive pulmonary disease, multiple sclerosis |
| **2** | 3 red points for use of recreational drugs such as methamphetamine or opioids |
| **3** | 2 red points when groups were not matched for age |
| **4** | 2 red points when groups were not matched for sex |
| **5** | 2 red points for medication use as for example immunomodulators |
| **6** | 2 red points for early traumatic life events |
| **7** | 2 red points for shift work and primary sleep disorders |
| **8** | 1.5 red points for use of antipsychotics |
| **9** | 1 red point for more common systemic immune disorders including diabetes type 1/2, essential hypertension, metabolic syndrome |
| **10** | 1 red point for not fasting (8 hours before blood extraction) |
| **11** | 1 red point for use of omega-3 and antioxidant supplements |
| **12** | 1 red point when data were not controlled for body mass index |
| **13** | 1 red point when data were not controlled for physical activity or sedentary life |
| **14** | 1 red point when data were not controlled for smoking |

| | |
|---|---|
| **15** | 1 red point for use of oral contraceptives or NSAIDs |
| **16** | 0.5 red points when data were not controlled for ethnicity in countries such as US, Brazil |
| **17** | 0.5 red points when data were not controlled for seasonality |
| **18** | 0.5 red points when data were not controlled for diurnal variation (8-10 a.m. versus all other time points) |
| | **Total red point score (26 points)** |

**ESF, Table 3.** PRISMA checklist

| Section/topic | # | Checklist item | Reported on page # |
|---|---|---|---|
| **TITLE** | | | |
| Title | 1 | Identify the report as a systematic review, meta-analysis, or both. | 1 |
| **ABSTRACT** | | | |
| Structured summary | 2 | Provide a structured summary including, as applicable: background; objectives; data sources; study eligibility criteria, participants, and interventions; study appraisal and synthesis methods; results; limitations; conclusions and implications of key findings; systematic review registration number. | 3 |
| **INTRODUCTION** | | | |
| Rationale | 3 | Describe the rationale for the review in the context of what is already known. | 5-7 |
| Objectives | 4 | Provide an explicit statement of questions being addressed with reference to participants, interventions, comparisons, outcomes, and study design (PICOS). | 8 |
| **METHODS** | | | |
| Protocol and registration | 5 | Indicate if a review protocol exists, if and where it can be accessed (e.g., Web address), and, if available, provide registration information including registration number. | 9 |
| Eligibility criteria | 6 | Specify study characteristics (e.g., PICOS, length of follow-up) and report characteristics (e.g., years considered, language, publication status) used as criteria for eligibility, giving rationale. | 10 |
| Information sources | 7 | Describe all information sources (e.g., databases with dates of coverage, contact with study authors to identify additional studies) in the search and date last searched. | 11 |
| Search | 8 | Present full electronic search strategy for at least one database, including any limits used, such that it could be repeated. | 9-10, ESF, table 1 |
| Study selection | 9 | State the process for selecting studies (i.e., screening, eligibility, included in systematic review, and, if applicable, included in the meta-analysis). | 10-11, Figure 2 |

| | | | |
|---|---|---|---|
| Data collection process | 10 | Describe method of data extraction from reports (e.g., piloted forms, independently, in duplicate) and any processes for obtaining and confirming data from investigators. | 11 |
| Data items | 11 | List and define all variables for which data were sought (e.g., PICOS, funding sources) and any assumptions and simplifications made. | 11 |
| Risk of bias in individual studies | 12 | Describe methods used for assessing risk of bias of individual studies (including specification of whether this was done at the study or outcome level), and how this information is to be used in any data synthesis. | 12-13 |
| Summary measures | 13 | State the principal summary measures (e.g., risk ratio, difference in means). | 12-13 |
| Synthesis of results | 14 | Describe the methods of handling data and combining results of studies, if done, including measures of consistency (e.g., $I^2$) for each meta-analysis. | 12-13 |
| Risk of bias across studies | 15 | Specify any assessment of risk of bias that may affect the cumulative evidence (e.g., publication bias, selective reporting within studies). | 12-13 |
| Additional analyses | 16 | Describe methods of additional analyses (e.g., sensitivity or subgroup analyses, meta-regression), if done, indicating which were pre-specified. | 12-13 |
| **RESULTS** | | | |
| Study selection | 17 | Give numbers of studies screened, assessed for eligibility, and included in the review, with reasons for exclusions at each stage, ideally with a flow diagram. | 14 |
| Study characteristics | 18 | For each study, present characteristics for which data were extracted (e.g., study size, PICOS, follow-up period) and provide the citations. | 15, ESF, table 4 |
| Risk of bias within studies | 19 | Present data on risk of bias of each study and, if available, any outcome level assessment (see item 12). | Table 3 |
| Results of individual studies | 20 | For all outcomes considered (benefits or harms), present, for each study: (a) simple summary data for each intervention group (b) effect estimates and confidence intervals, ideally with a forest plot. | Table 1 |
| Synthesis of results | 21 | Present results of each meta-analysis done, including confidence intervals and measures of consistency. | Table 2 |
| Risk of bias across studies | 22 | Present results of any assessment of risk of bias across studies (see Item 15). | Table 3 |
| Additional analysis | 23 | Give results of additional analyses, if done (e.g., sensitivity or subgroup analyses, meta-regression [see Item 16]). | 18, ESF, table 5 |

| | | | |
|---|---|---|---|
| **DISCUSSION** | | | |
| Summary of evidence | 24 | Summarize the main findings including the strength of evidence for each main outcome; consider their relevance to key groups (e.g., healthcare providers, users, and policy makers). | 19-26 |
| Limitations | 25 | Discuss limitations at study and outcome level (e.g., risk of bias), and at review-level (e.g., incomplete retrieval of identified research, reporting bias). | 26 |
| Conclusions | 26 | Provide a general interpretation of the results in the context of other evidence, and implications for future research. | 26 |
| **FUNDING** | | | |
| Funding | 27 | Describe sources of funding for the systematic review and other support (e.g., supply of data); role of funders for the systematic review. | 27 |

**ESF, Table 4.** Characteristics of the studies included in the systematic reviews and meta-analysis

| NO | Authors, years | Setting | Type of case | Type of Control | Sample Size | | | Age | | Assessed Biomarkers | Specimen | Method | Quality score | Red point score | Findnigs |
|---|---|---|---|---|---|---|---|---|---|---|---|---|---|---|---|
| | | | | | Cases M/F | Control M/F | Total M/F | Case-Mean (SD) | Control-Mean(SD) | | | | | | |
| 1 | Aarsland, Leskauskaite et al. 2019 | Norway | MDD-Psychotic feature | Healthy Control | 27 12/15 | 14 6/8 | 41 18/23 | 45,3(10,7) | - | TRP,KYN, KA,3HK, XA, AA, 3HA,QA,PA | Serum | LCMS | 5,75 | 13,5 | TRP*,KYN#, KA*,3HK*, XA*, AA*, 3HA*,QA#,PA* |
| 2 | Achtyes, Keaton et al. 2020 | USA | Post partum depression-Suicidal | Healthy Control | 87 0/87 | 60 0/60 | 147 0/147 | 26.7(5.2) | 28(6.3) | TRP, KYN, KA,QA | Plasma | HPLC-UV | 5,75 | 14 | TRP*, KYN*, KA*,QA* |
| 3 | van den Ameele, van Nuijs et al. 2020 | Belgium | BD-Depression-Psychotic feature | Healthy Control | 35 11/24 | 35 16/29 | 70 27/53 | 43.7(9.7) | 42.7(11.6) | TRP, KYN, 3HK,QA, KA | Plasma | UPLC-MS/MS | 7,75 | 10 | TRP*, KYN*, 3HK*,QA*, KA* |
| | | | BD-hypomania-Psychotic feature | Healthy Control | 32 15/17 | 35 16/29 | 67 31/46 | 42.4(12.7) | 42.7(11.6) | TRP, KYN, 3HK,QA, KA | Plasma | UPLC-MS/MS | | | TRP*, KYN*, 3HK*,QA*, KA* |
| 4 | Anderson, Parry-Billings et al. 1990 | UK | MDD-Melancholia | Healthy Control | 32 15/16 | 31 15/16 | 62 30/32 | 34.4(8.5) | 33.9(7.7) | TRP | Plasma | Fluorometry | 4 | 12,5 | TRP* |
| 5 | Bay-Richter, Linderholm et al. 2015 | Sweden | Affective disorders-Suicidal | Healthy Control | 30 9/21 | 36 29/7 | 66 38/28 | 40(8,7) | 29,7(7,3) | QA,KA | CSF | GCMS,HPLC | 3 | 18,5 | QA#,KA* |
| 6 | Bradley, Case et al. 2015 | USA | MDD-Non suicidal | Healthy Control | 30 18/12 | 22 9/13 | 52 27/25 | 15.2(1.9) | 15.9(2.6) | TRP. KYN, 3HA | Plasma | HPLC | 4,25 | 16 | TRP#. KYN*, 3HA# |
| | | | MDD-suicidal | Healthy Control | 20 5/15 | 22 9/13 | 42 14/28 | 16.8(1.8) | 15.9(2.6) | TRP. KYN, 3HA | Plasma | HPLC | | | TRP*. KYN*, 3HA* |
| 7 | Brundin, Sellgren et al. 2016 | Sweden | MDD-Suicidal | Healthy Control | 22 - | 36 - | 58 - | 42.1(13.6) | 32.5(11.3) | PA | CSF | GCMS | 5,5 | 18 | PA* |
| | | | Dysthymia | Healthy Control | 4 - | 36 - | 40 - | 44.3(23.3) | 32.5(11.3) | PA | CSF | GCMS | | | PA* |
| | | | Depression NOS | Healthy Control | 9 - | 36 - | 45 - | 30.8(6.6) | 32.5(11.3) | PA | CSF | GCMS | | | PA* |
| | | | MDD-Suicidal | Healthy Control | 15 - | 29 - | 44 - | 43.1(13.5) | 40.6(11.5) | PA, QA | Plasma | GCMS | | | PA*, QA# |
| | | | Dysthymia | Healthy Control | 3 - | 29 - | 32 - | 43.0(18.9) | 40.6(11.5) | PA, QA | Plasma | GCMS | | | PA*, QA* |
| | | | BD | Healthy Control | 21 - | 29 - | 50 - | 39.2(12.4) | 40.6(11.5) | PA, QA | Plasma | GCMS | | | PA*, QA# |
| | | | Depression UNS | Healthy Control | 4 - | 29 - | 33 - | 28.1(8.5) | 40.6(11.5) | PA, QA | Plasma | GCMS | | | PA*, QA# |

| | | | | | | | | | | | | | | | |
|---|---|---|---|---|---|---|---|---|---|---|---|---|---|---|---|
| 8 | Busse, Busse et al. 2015 | Germany | Depression-Suicidal | Healthy Control | 6<br>2/4 | 10<br>5/5 | 16<br>7/9 | 47(10) | 56(6) | QA | Brain Tissue | - | 2,75 | 16 | QA* |
| 9 | Clark, Pocivavsek et al. 2016 | USA | Depression-Suicidal | Healthy Control | 45<br>30/15 | 36<br>27/9 | 91<br>57/24 | 43.3(13.6) | 42.1(14.3) | TRP,KYN, KA,3-HK, QA | Brain Tissue | HPLC,GC/MS | 2,75 | 17,5 | TRP*,KYN*, KA*,3-HK*, QA* |
| 10 | Cowen, Parry-Billings et al. 1989 | UK | MDD-Melancholia | Healthy Control | 12<br>6/6 | 12<br>6/6 | 24<br>12/12 | 36,5<br>(26-55) | 36,2<br>(28-54) | TRP | Plasma | Fluorometry | 6,75 | 11 | TRP* |
| 11 | Dahl, Andreassen et al. 2015 | Norway | MDD-Melancholia | Healthy Control | 50<br>12/38 | 34<br>15/19 | 84<br>27/57 | 40(12) | 38,3(13,9) | TRP,KYN, KA, QA | Plasma | HPLC | 4,25 | 15 | TRP$^{\#}$,KYN*, KA$^{\#}$, QA* |
| 12 | Erhardt, Lim et al. 2013 | Sweden | MDD-Suicidal | Healthy Control | 29 | 7 | 36 | - | 30(18-66) | KA, QA | CSF | HPLC-GCMS | 4,5 | 15 | KA*, QA$^{\#}$ |
| 13 | Gabbay, Klein et al. 2010 | USA | MDD-Melancholia | Healthy Control | 20<br>9/11 | 22<br>9/13 | 42<br>18/24 | 15.6(2.0) | 16.0(2.7) | TRP, KYN | Plasma | HPLC | 6,25 | 9,5 | TRP*, KYN* |
| | | | MDD-non Melancholia | | 30<br>14/16 | | 52<br>23/29 | 16.3(2.1) | | | | | | | TRP#, KYN* |
| 14 | Hoekstra, Fekkes et al. 2006 | Netherland | BD-Psychotic feature | Healthy Control | 20<br>14/6 | 20<br>14/6 | 40<br>28/12 | 50,2(13,8) | 50,1(13,5) | TRP | Plasma | HPLC | 3,75 | 15 | TRP* |
| 15 | Maes, Jacobs et al. 1990 | Belgium | MDD-Melancholia | Healthy Control | 15<br>6/9 | 16<br>8/8 | 31<br>14/17 | 41.0(15.8) | 40.1(10.1) | TRP | Plasma | LC | 6 | 10,5 | TRP* |
| | | | | | 22<br>10/12 | | 38<br>18/20 | 43.9(13.8) | | | | | | | TRP* |
| | | | | | 13<br>6/7 | | 29<br>14/15 | 52.2(14.5) | | | | | | | TRP* |
| 16 | Maes, Meltzer et al. 1993 | Belgium | MDD-Melancholia | Healthy Control | 7 | 8 | 15 | 33.0(13.1) | 44.1(13.1) | TRP | Plasma | LC | 6,75 | 9,5 | TRP* |
| | | | | | 7 | 8 | 15 | 51.7(17.0) | | | | | | | TRP* |
| | | | | | 10 | 8 | 18 | 54.8(12.5) | | | | | | | TRP* |
| 17 | Maes, De Backer et al. 1995 | Belgium | MDD-Melancholia | Healthy Control | 41<br>17/24 | 50<br>26/24 | 91<br>43/48 | 42.1(14.2) | 40.7(11.3) | TRP | Plasma | LC | 6,25 | 12 | TRP* |
| | | | | | 47<br>12/35 | | 97<br>38/59 | 47.3(13.5) | | | | | | | TRP* |
| | | | | | 35<br>14/21 | | 85<br>40/45 | 51.7(15.0) | | | | | | | TRP* |
| 18 | Maes, Wauters et al. 1996 | Belgium | MDD-Melancholia | Healthy Control | 12<br>4/8 | 24<br>11/13 | 36<br>15/21 | 42,8(11,9) | 41,1(13,5) | TRP | Serum | HPLC | 5,75 | 11,5 | TRP* |
| | | | | | 21<br>3/18 | | 45<br>14/31 | 48,7(14,3) | | | | | | | TRP* |
| | | | | | 9<br>3/6 | | 33<br>14/19 | 58,7(14) | | | | | | | TRP* |
| 19 | Milaneschi, Allers et al. 2021 | Netherland | MDD-remitted-Melancholia | Healthy Control | 753<br>226/527 | 642<br>246/396 | 1395<br>472/923 | 43,5(12,5) | 41,1(14,7) | TRP,KYN, KA, QA | Plasma | - | 3 | 17,5 | TRP*,KYN*, KA*, QA* |
| | | | MDD-current-Melancholia | | 1100<br>358/743 | | 1742<br>603/1139 | 40,8(12,1) | | | | | | | TRP$^{\#}$,KYN*, KA*, QA* |

| | | | | | | | | | | | | | | |
|---|---|---|---|---|---|---|---|---|---|---|---|---|---|---|
| 20 | Miller, Llenos et al. 2006 | USA | MDD-Psychotic feature | Healthy Control | 14 | 14 | 28 | 45.9(9.7) | 48.6(10.8) | KYN, KA | Brain Tissue | HPLC | 4,75 | 12,5 | KYN$^{\#}$, KA$^{\#}$ |
| | | | BD-Psychotic feature | | 14 | 14 | 28 | 41.8(11.9) | | | | | | | KYN$^{\#}$, KA$^{\#}$ |
| 21 | Møller 1993 | Denmark | Depression-Melancholia | Healthy Control | 26<br>8/18 | 55<br>16/39 | 81<br>24/57 | 52(10) | 38(13) | TRP | Plasma | IEC | 3,25 | 17 | TRP* |
| 22 | Myint, Kim et al. 2007a | South Korea | BD-Psychotic feature | Healthy Control | 39<br>15/24 | 80<br>40/40 | 119<br>55/64 | 37.6(11.6) | 39.0(8.7) | TRP, KYN, KA, 3HA | Plasma | HPLC | 6 | 12 | TRP*, KYN*, KA*, 3HA$^{\#}$ |
| 23 | Myint, Kim et al. 2007b | South Korea | Depression-Melancholia | Healthy Control | 58<br>48/10 | 189<br>32/157 | 247<br>80/167 | 44.6(14.6) | 32.4(10.6) | TRP, KYN, KA, 3HA | Plasma | HPLC | 5,75 | 15 | TRP*, KYN*, KA*, 3HA$^{\#}$ |
| 24 | Pompili, Lionetto et al. 2019 | Italy | MDD-Suicidal | Healthy Control | 49<br>24/25 | 78<br>34/44 | 127<br>58/69 | 36,6(8,5) | 40,4(12,3) | TRP, KYN AA, XA | Plasma | LCMS | 3 | 18,5 | TRP*, KYN AA, XA |
| 25 | Price, Charney et al. 1991 | USA | Depression-Melancholia and psychotic feature | Healthy Control | 126<br>38/88 | 58<br>17/41 | 184<br>58/126 | 43(14) | 38(13) | TRP | Serum | HPLC | 7 | 15 | TRP* |
| 26 | Quintana 1992 | Spain | MDD-Melancholia | Healthy Control | 25<br>10/15 | 25<br>10/15 | 50<br>20/30 | 54(9,3) | - | TRP | Plasma | - | 5,75 | 13,5 | TRP* |
| 27 | Ryan, Allers et al. 2020 | Ireland | Depression-Psychotic feature | Healthy Control | 94<br>36/58 | 57<br>20/37 | 151<br>56/95 | 55.4(14.7) | 51.7(11.4) | TRP, KYN KA, AA, 3HK, XA 3HA, QA PA | Plasma | LC/MS | 6 | 13,5 | TRP*, KYN* KA*, AA*, 3HK*, XA* 3HA*, QA* PA* |
| 28 | Savitz, Drevets et al. 2015d | USA | MDD-Suicidal | Healthy Control | 53<br>11/42 | 47<br>18/29 | 100<br>29/71 | 34.6(9.8) | 34.3(11.4) | TRP, KYN KA, 3HK, QA | Serum | LC–MS/MS | 7 | 11,5 | TRP*, KYN* KA*, 3HK$^{\#}$, QA$^{\#}$ |
| 29 | Sellgren, Gracias et al. 2019 | Sweden | BD-Psychotic feature | Healthy Control | 163<br>64/99 | 114<br>52/62 | 277<br>116/161 | 34.2(3.3) | 35.1(3.1) | KA | Plasma | HPLC | 6 | 13,5 | KA* |
| | | | | | 94<br>39/55 | 113<br>51/62 | 207<br>90/117 | 36(20.5) | - | | CSF | | | | KA$^{\#}$ |
| 30 | Song, Lin et al. 1998 | Belgium | Depression-Melancholia | Healthy Control | 6<br>2/4 | 14<br>6/8 | 20<br>8/12 | 50.3(15.3) | 45.5(15.5) | TRP | Plasma | HPLC | 4,25 | 13,5 | TRP* |
| 31 | Steiner, Walter et al. 2011 | Germany | Depression-Suicidal | Healthy Control | 12<br>6/6 | 10<br>5/5 | 22<br>11/11 | 51(9) | 56(6) | QA | Brain Tissue | ELISA | 3,75 | 16 | QA$^{\#}$ |
| 32 | Sublette, Galfalvy et al. 2011 | USA | MDD-Suicidal | Healthy Control | 30<br>16/14 | 31<br>10/21 | 61<br>26/35 | 37.8(13.08 | 35.6(13.9) | TRP, KYN | Serum | HPLC | 4 | 14 | TRP*, KYN$^{\#}$ |
| 33 | Trepci, Sellgren et al. 2021 | Sweden | BD-Suicidal | Healthy Control | 101<br>40/61 | 80<br>39/41 | 181<br>79/102 | 44.2(13.9) | 34.7(12.4) | TRP, KYN KA, PA, QA | CSF | UPLC-MS/MS | 7 | 16,5 | TRP*, KYN$^{\#}$ KA$^{\#}$, PA$^{\#}$, QA$^{\#}$ |
| 34 | Zhou, Zheng et al. 2018 | China | MDD-Suicidal | Healthy Control | 84<br>39/45 | 60<br>35/25 | 144<br>74/70 | 35.0(12.1) | 31.6(10.7) | TRP,KYN, KA | Serum | HPLC-MS/MS | 5 | 12,5 | TRP*,KYN*, KA* |

| | | | | | | | | | | | | | | | |
|---|---|---|---|---|---|---|---|---|---|---|---|---|---|---|---|
| 35 | Messaoud, Mensi et al. 2019 | Tunis | MDD | Healthy Control | 56 20/36 | 40 15/25 | 96 35/61 | 34.2(11.3) | 38.7(9.2) | TRP, KYN | Plasma | ECLIA | 6 | 10 | TRP*, KYN# |

*: Indicates that patients have reduced level of the measured metabolite compared to healthy control

#: Indicates that patients have increased level of the measured metabolites compared to healthy control

TRP: Tryptophan, KYN: Kynurenine, KA: Kynurenic acid, 3HK: 3-Hydroxykynurenine, AA: Anthranilic acid, 3HA: 3-Hydroxyanthranilic acid, XA: Xanthurenic acid, QA: Quinolinic acid, PA: Picolinic acid, MDD: Major depressive disorder, BD: Bipolar disorder, MDD-M: Major depressive disorder with melancholia, DD: Dysthymic disorder, UD: Unipolar depression, HPLC: High performance liquid chromatography, HPLC-MS/MS: High performance liquid chromatography with tandem mass spectrometry, CE-TOFMS: Capillary Electrophoresis Time Of Flight Mass Spectrometer, LC-MS/MS: Liquid chromatography with tandem mass spectrometry, UPLC-MS/MS: Ultra performance liquid chromatography with tandem mass spectrometry, LCECA: Liquid, chromatography electrochemical array, XLC-MS/MS: Extraction-liquid chromatographic-tandem mass spectrometric, GCMS: Gas chromatography–mass spectrometry, LC: Liquid chromatography, HPLC-UV: High perfomance liquid chromatography- Ultra-violate

**ESF. Table 5**. Results of Meta-regression

| Variables | No. of Studies | Covariates | 1-sided p-value | Z-Value |
|---|---|---|---|---|
| TRP | 29 | Plasma | 0.030 | -1.88 |
| TRP/CAAs | 7 | ECT-No | 0.001 | -3.10 |
| CAAs | 5 | Sample size | 0.042 | 1.72 |
| KYN | 17 | Plasma | 0.007 | -2.44 |
| KA/KYN | 9 | Male gender | 0.048 | -1.66 |
| | | Female gender | 0.038 | 1.77 |
| (KYN+3HK+3HA+XA+QA+PA) | 26 | Plasma | 0.003 | -2.75 |
| KA | 13 | Latitude | 0.015 | 2.16 |
| | | Plasma | 0.026 | -1.94 |
| | 12 | Partially Medicated | 0.049 | -1.65 |
| QA | 14 | Latitude | 0.004 | 2.64 |
| | | Plasma | 0.014 | -2.17 |
| | 13 | Partially Medicated | 0.000 | 4.76 |

TRP: Tryptophan, KYN: Kynurenine, KA: Kynurenic acid, 3HK: 3-Hydroxykynurenine, AA: Anthranilic acid, 3HA: 3-Hydroxyanthranilic acid, XA: Xanthurenic acid, QA: Quinolinic acid, PA: Picolinic acid, MDD: CAAs: Competing amino acids.

#### CAAs

| Study name | Year | Std diff in means | p-Value | Sample size: Patients | Sample size: Control |
|---|---|---|---|---|---|
| Anderson-M | 1990 | 0,002 | 0,994 | 16 | 31 |
| Cowen-M | 1989 | -0,767 | 0,018 | 12 | 58 |
| Maes-a-M | 1990 | -0,049 | 0,895 | 13 | 16 |
| Maes-b-M | 1993 | -1,064 | 0,036 | 10 | 8 |
| Maes-c-M | 1995 | -0,080 | 0,716 | 35 | 50 |
| | | -0,304 | 0,107 | 86 | 163 |

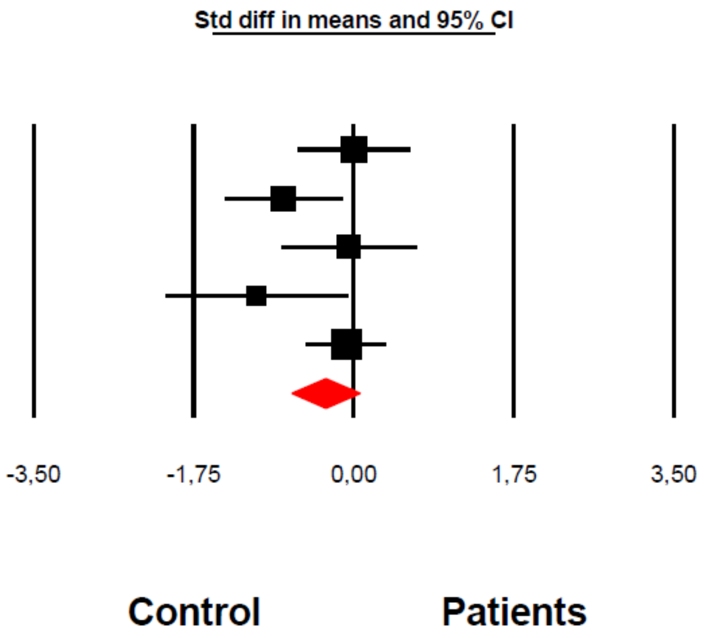

**Almulla et al, 2022**

**ESF, Figure 1**: The forest plot of competing amino acids (CAAs) level between severe affective disorder patients and healthy control.

#### TRP/CAAs

| Study name | Year | Std diff in means | p-Value | Patients | Control |
|---|---|---|---|---|---|
| Cowen-M | 1989 | -0,377 | 0,346 | 12 | 35 |
| Anderson-M | 1990 | -0,551 | 0,083 | 16 | 31 |
| Maes-a-M | 1990 | -0,576 | 0,139 | 13 | 16 |
| Maes-b-M | 1993 | -0,340 | 0,523 | 10 | 8 |
| Moller-a-M | 1993 | -0,595 | 0,032 | 18 | 55 |
| Maes-c-M | 1995 | -0,335 | 0,135 | 35 | 50 |
| Maes-e-M | 1996 | -1,937 | 0,000 | 9 | 24 |
| | | -0,617 | 0,000 | 113 | 219 |

Sample size: Patients, Control

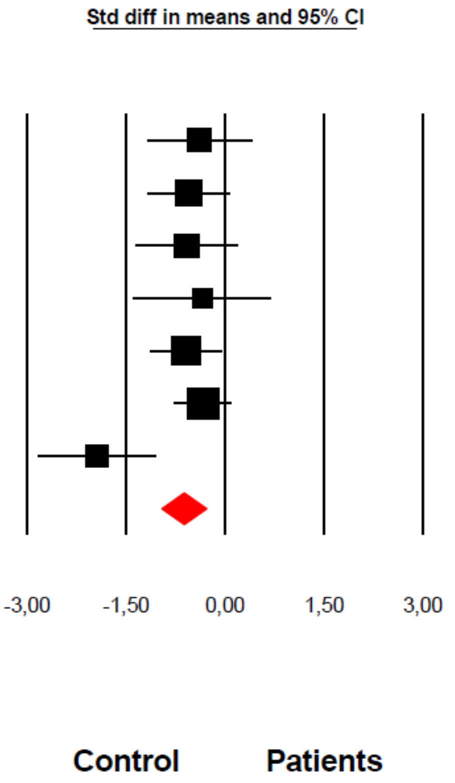

Almulla et al, 2022

**ESF, Figure 2**: The forest plot of TRP/CAAs ratio between severe affective disorder patients and healthy control.

#### KYN

| Study name | Year | Std diff in means | p-Value | Sample size: Patients | Sample size: Control |
|---|---|---|---|---|---|
| Aarsland-P | 2019 | 0,320 | 0,334 | 27 | 14 |
| Achtyes-S | 2020 | -0,442 | 0,009 | 87 | 60 |
| Ameele-P | 2020 | -0,448 | 0,067 | 34 | 35 |
| Bradley-S | 2015 | -0,662 | 0,037 | 20 | 22 |
| Clark-S | 2016 | -0,119 | 0,594 | 45 | 36 |
| Dahl-M | 2015 | -0,281 | 0,237 | 38 | 34 |
| Gabbay-M | 2010 | -0,224 | 0,469 | 20 | 22 |
| Milaneschi-M | 2021 | -0,200 | 0,000 | 927 | 642 |
| Myint-a-P | 2007 | -0,302 | 0,123 | 39 | 80 |
| Myint-b-S | 2007 | -0,129 | 0,389 | 58 | 189 |
| Pompili-S | 2018 | -1,659 | 0,000 | 49 | 78 |
| Ryan-P | 2020 | -0,089 | 0,596 | 94 | 57 |
| Savitz-c-S | 2015 | 0,044 | 0,846 | 35 | 58 |
| Sublette-S | 2011 | 0,883 | 0,008 | 14 | 31 |
| Trepci-S | 2021 | 0,739 | 0,000 | 101 | 80 |
| Zhou-a-S | 2018 | -0,045 | 0,790 | 84 | 60 |
| Messaoud-S | 2018 | 0,813 | 0,000 | 56 | 40 |
| | | -0,114 | 0,350 | 1727 | 1538 |

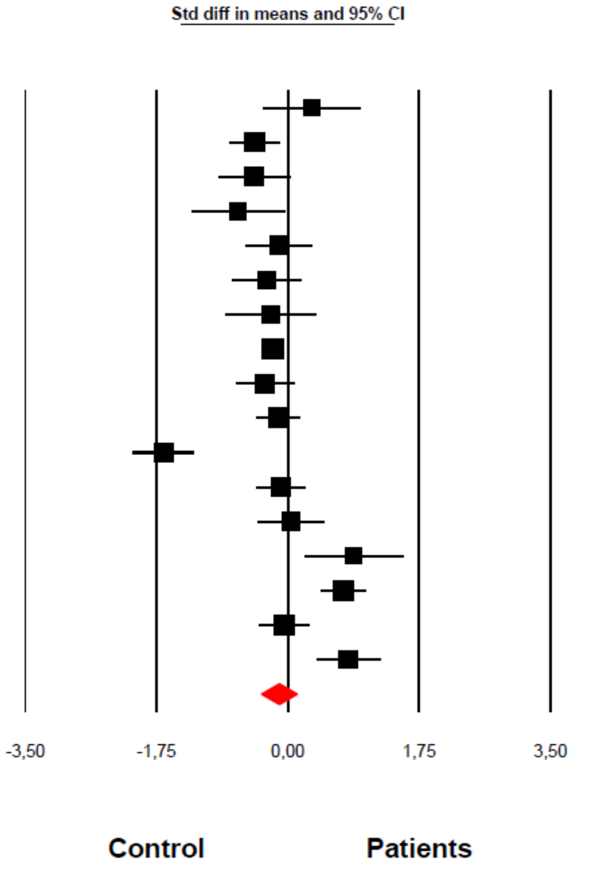

**Almulla et al, 2022**

**ESF, Figure 3**: The forest plot of kynurenine (KYN) level between severe affective disorder patients and healthy control.

### (KYN+3HK+3HA+XA+QA+PA)

| Study name | Year | Std diff in means | p-Value | Sample size Patients | Sample size Control |
|---|---|---|---|---|---|
| Aarsland-P | 2019 | -0,299 | 0,371 | 27 | 14 |
| Myint-a-P | 2007 | -0,103 | 0,600 | 39 | 80 |
| Myint-b-S | 2007 | -0,041 | 0,785 | 58 | 189 |
| Ryan-P | 2020 | -0,266 | 0,118 | 94 | 57 |
| Bradley-S | 2015 | -0,667 | 0,036 | 20 | 22 |
| Gabbay-M | 2010 | -0,256 | 0,410 | 20 | 22 |
| Ameele-P | 2020 | -0,448 | 0,067 | 34 | 35 |
| Clark-S | 2016 | -0,546 | 0,018 | 45 | 36 |
| Savitz-c-S | 2015 | 0,069 | 0,761 | 35 | 58 |
| Pompili-S | 2018 | -0,920 | 0,000 | 49 | 78 |
| Achtyes-S | 2020 | -0,402 | 0,018 | 87 | 60 |
| Bay-Richte-S | 2015 | 3,244 | 0,000 | 30 | 36 |
| Dahl-M | 2015 | -0,189 | 0,424 | 38 | 34 |
| Erhardt-S | 2013 | 0,967 | 0,000 | 31 | 36 |
| Miller-P | 2006 | 0,253 | 0,505 | 14 | 14 |
| Miller-P | 2006 | 0,187 | 0,621 | 14 | 14 |
| Trepci-S | 2021 | 0,539 | 0,000 | 101 | 80 |
| Zhou-a-S | 2018 | -0,045 | 0,790 | 84 | 60 |
| Milaneschi-M | 2021 | -0,131 | 0,011 | 927 | 642 |
| Sublette-S | 2011 | 0,883 | 0,008 | 14 | 31 |
| Brundin-CeS | 2016 | -2,611 | 0,000 | 12 | 36 |
| Brundin-PeS | 2016 | -0,223 | 0,445 | 21 | 29 |
| Brundin-PeS | 2016 | -0,350 | 0,490 | 7 | 29 |
| Busse-S | 2015 | 1,459 | 0,012 | 6 | 10 |
| Steiner-S | 2011 | 0,828 | 0,067 | 12 | 10 |
| Messaoud-S | 2018 | 0,813 | 0,000 | 56 | 40 |
| | | 0,048 | 0,692 | 1874 | 1752 |

Std diff in means and 95% CI

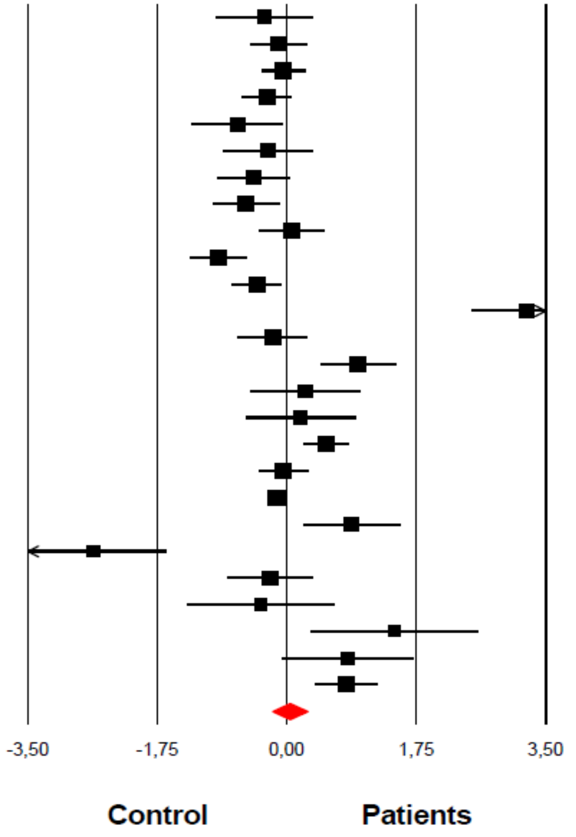

**Almulla et al, 2022**

**ESF, Figure 4**: The forest plot of neurotoxic composite (KYN+3HK+3HA+XA+QA+PA) level between severe affective disorder patients and healthy control.

#### KA/KYN

| Study name | Year | Subgroup within study | Statistics for each study: Std diff in means | p-Value | Sample size: Patients | Control |
|---|---|---|---|---|---|---|
| Dahl-M | 2015 | Melancholic | 0,196 | 0,408 | 38 | 34 |
| Milaneschi-M | 2021 | Melancholic | 0,042 | 0,417 | 927 | 642 |
| | | Melancholic | 0,049 | 0,333 | 965 | 676 |
| Aarsland-P | 2019 | Psychotic | -0,399 | 0,230 | 27 | 14 |
| Miller-P | 2006 | Psychotic | -0,063 | 0,867 | 14 | 14 |
| Miller-P | 2006 | Psychotic | -0,063 | 0,868 | 14 | 14 |
| Myint-a-P | 2007 | Psychotic | -0,123 | 0,533 | 39 | 80 |
| Ryan-P | 2020 | Psychotic | -0,264 | 0,121 | 94 | 57 |
| | | Psychotic | -0,201 | 0,066 | 188 | 179 |
| Achtyes-S | 2020 | Suicidal | 0,061 | 0,718 | 87 | 60 |
| Clark-S | 2016 | Suicidal | -0,153 | 0,496 | 45 | 36 |
| Messaoud-S | 2018 | Suicidal | -0,813 | 0,000 | 56 | 40 |
| Myint-b-S | 2007 | Suicidal | -0,407 | 0,008 | 58 | 189 |
| Savitz-c-S | 2015 | Suicidal | -0,174 | 0,443 | 35 | 58 |
| Trepci-S | 2021 | Suicidal | -0,070 | 0,649 | 101 | 80 |
| Zhou-a-S | 2018 | Suicidal | -0,147 | 0,385 | 84 | 60 |
| | | Suicidal | -0,231 | 0,024 | 466 | 523 |
| | | Overall | -0,035 | 0,410 | 1619 | 1378 |

Std diff in means and 95% CI

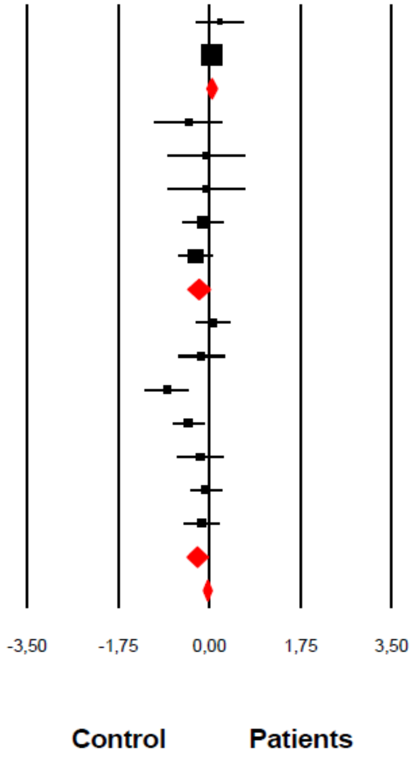

Almulla et al, 2022

**ESF, Figure 5**: The forest plot of kynurenic acid (KA)/kynurenine (KYN) ratio between severe affective disorder patients and healthy control.

# KA

| Study name | | Std diff in means | p-Value | Sample size: Patients | Sample size: Control |
|---|---|---|---|---|---|
| Aarsland-P | 2019 | -0,478 | 0,152 | 27 | 14 |
| Achtyes-S | 2020 | -0,319 | 0,059 | 87 | 60 |
| Clark-S | 2016 | -0,425 | 0,060 | 45 | 36 |
| Dahl-M | 2015 | 0,111 | 0,640 | 38 | 34 |
| Milaneschi-M | 2021 | -0,116 | 0,025 | 927 | 642 |
| Miller-P | 2006 | 0,126 | 0,739 | 14 | 14 |
| Miller-P | 2006 | 0,062 | 0,870 | 14 | 14 |
| Myint-a-P | 2007 | -0,548 | 0,006 | 39 | 80 |
| Myint-b-S | 2007 | -0,943 | 0,000 | 58 | 189 |
| Ryan-P | 2020 | -0,616 | 0,000 | 94 | 57 |
| Savitz-c-S | 2015 | -0,304 | 0,181 | 35 | 58 |
| Trepci-S | 2021 | 0,599 | 0,000 | 101 | 80 |
| Zhou-a-S | 2018 | -0,339 | 0,046 | 84 | 60 |
| | | -0,260 | 0,024 | 1563 | 1338 |

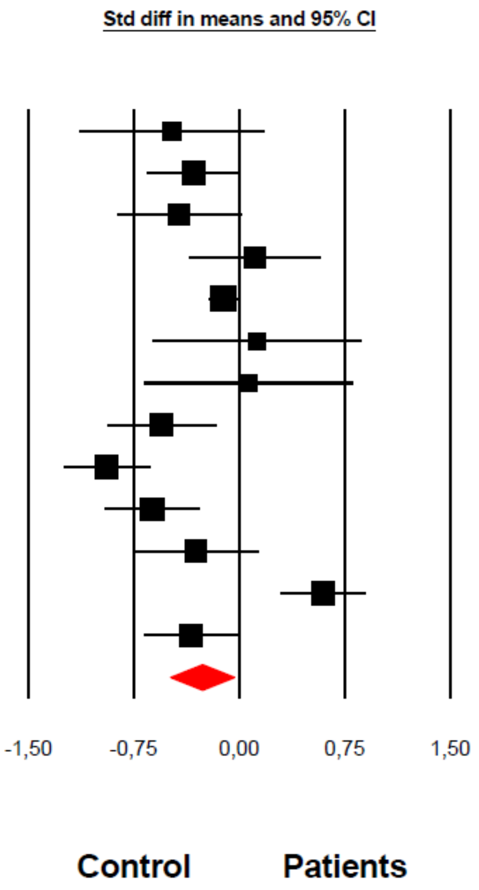

Almulla et al, 2022

**ESF, Figure 6**: The forest plot of kynurenic acid (KA) level between severe affective disorder patients and healthy control.

# AA

| Study name | Year | Std diff in means | p-Value | Sample size: Patients | Sample size: Control |
|---|---|---|---|---|---|
| Aarsland-P | 2019 | -0,472 | 0,157 | 27 | 14 |
| Pompili-S | 2018 | -0,047 | 0,796 | 49 | 78 |
| Ryan-P | 2020 | -0,362 | 0,033 | 94 | 57 |
| | | -0,248 | 0,040 | 170 | 149 |

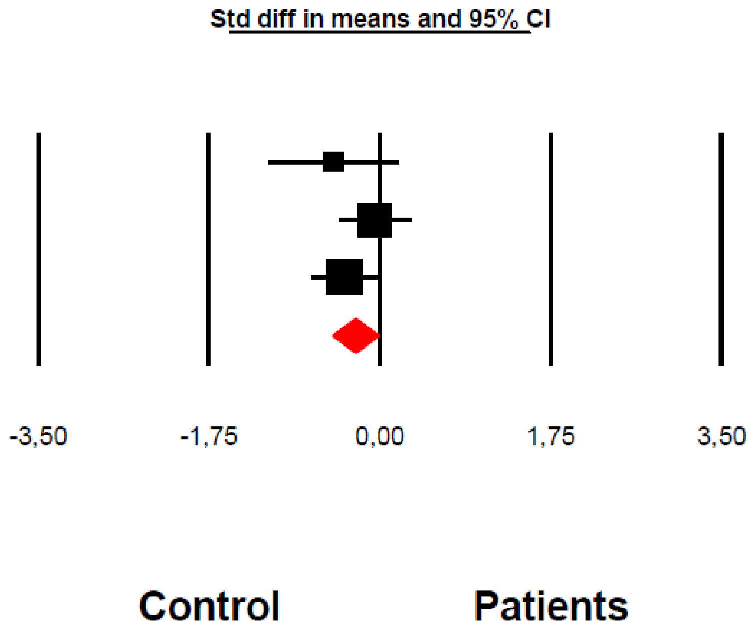

**Almulla et al, 2022**

**ESF, Figure 7:** The forest plot of anthranilic acid (AA) level between severe affective disorder patients and healthy control.

# QA

| Study name | Year | Std diff in means | p-Value | Sample size Patients | Sample size Control |
|---|---|---|---|---|---|
| Steiner-S | 2011 | 0,828 | 0,067 | 12 | 10 |
| Erhardt-S | 2013 | 0,967 | 0,000 | 31 | 36 |
| Savitz-c-S | 2015 | 0,204 | 0,369 | 35 | 58 |
| Bay-Richte-S | 2015 | 3,244 | 0,000 | 30 | 36 |
| Dahl-M | 2015 | -0,098 | 0,679 | 38 | 34 |
| Busse-S | 2015 | 1,459 | 0,012 | 6 | 10 |
| Clark-S | 2016 | -1,084 | 0,000 | 45 | 36 |
| Brundin-PeS | 2016 | 0,270 | 0,348 | 21 | 29 |
| Brundin-PeS | 2016 | 0,097 | 0,847 | 7 | 29 |
| Aarsland-P | 2019 | 0,077 | 0,815 | 27 | 14 |
| Ryan-P | 2020 | 0,174 | 0,301 | 94 | 57 |
| Achtyes-S | 2020 | -0,363 | 0,032 | 87 | 60 |
| Trepci-S | 2021 | 0,402 | 0,008 | 101 | 80 |
| Milaneschi-M | 2021 | -0,062 | 0,230 | 927 | 642 |
| | | 0,358 | 0,041 | 1461 | 1131 |

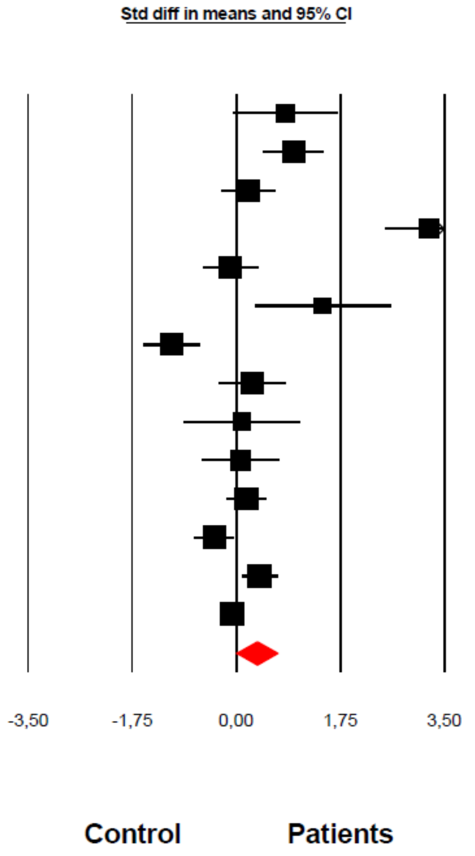

Almulla et al, 2022

**ESF, Figure 8:** The forest plot of quinolinic acid (QA) level between severe affective disorder patients and healthy control.

### Abbreviations:

TRP: Tryptophan

KYN: Kynurenine

KA: Kynurenic acid

3HK: 3-Hydroxykynurenine

AA: Anthranilic acid

3HA: 3-Hydroxyanthranilic acid

XA: Xanthurenic acid

QA: Quinolinic acid

PA: Picolinic acid

CAAs: Competing amino acids

IDO: Indoleamine 2,3 dioxygenase

TDO: Tryptophan 2,3 dioxygenase

KAT: Kynurenine aminotransferase

KMO: Kynurenine 3-monooxygenase

KYNU: Kynureninase

$NAD^+$: Nicotinamide adenine dinucleotide

TRYCATs: Tryptophan Catabolites

TRYCAT pathway: Tryptophan catabolite pathway

SMD: Standardized mean difference

IL: Interleukin

IFN-γ: Interferon-gamma

O&NS: Oxidative and nitrosative stress

NAD: Nicotinamide adenine dinucleotide

MOOSE: Meta-Analyses of Observational Studies in Epidemiology

CSF: Cerebrospinal fluid

SD: Standard deviation

IOR: Interquartile range

ICS: Immune confounder scale

CI: Confidence intervals

LC-MS: Liquid chromatography-mass spectrometry

LC-MS/MS: Liquid chromatography with two mass spectrometry

UHPLC-MS: Ultra-high-performance liquid-chromatography- mass spectrometry

LC-HRMS: Liquid chromatography–high-resolution mass spectrometry

LC-UV: Liquid chromatography-UV detection

NMDA: N-methyl-D-aspartate

ELICA: Electrochemiluminescence immunoassay
